# Auxiliary modalities add no detectable discrimination or calibration over structured EHR for 30-day COPD readmission: evidence from a leakage-audited MIMIC-IV cohort

**DOI:** 10.64898/2026.10.14.26365503

**Authors:** Marco Pozza, Nicolò Navarin, Silvia Gabrielli

## Abstract

**Background:** Hospital readmission is a common and costly outcome in patients with chronic obstructive pulmonary disease (COPD). Multimodal deep learning models combining structured electronic health record (EHR) data with physiological, imaging, and textual data have been proposed to improve its prediction, but whether such fusion outperforms EHR data alone, once temporal-leakage safeguards are applied, remains unclear.

**Methods:** Using a stratified MIMIC-IV cohort of COPD patients enriched to about 50% readmission (natural prevalence 18.1%; n=1,279; 895/192/192 training/validation/test), we predicted 30-day, all-cause readmission from four frozen, pre-trained encoders (electrocardiogram, chest radiograph, clinical notes, structured EHR). We audited and corrected temporal leakage in each modality’s admission-linkage pipeline, then compared three fusion architectures (naive concatenation, scalar gated fusion, EHR-anchored residual fusion) against unimodal baselines and an EHR-alone reference, using bootstrap significance testing, a missingness stress test, and calibration analysis with and without post-hoc recalibration.

**Results:** The audit corrected three previously undetected temporal-alignment issues, affecting 5–9% of patients for clinical notes, 29–43% for chest radiographs, and 53–58% for ECG. After correction, every modality except EHR showed near-chance discrimination alone (ROC-AUC 0.46–0.53), confirmed with a non-linear adapter probe. No fusion architecture significantly outperformed the EHR-alone baseline (ROC-AUC 0.63); removing auxiliary modalities improved rather than degraded fusion performance under stress testing. The raw Brier score was lower for all three fusion architectures than for EHR-alone (about 0.24 versus 0.32), but this reflected overconfidence of the EHR-alone probe (calibration slope 0.12); after logistic recalibration fitted on the validation set, Brier scores were indistinguishable (EHR-alone 0.239; fusion 0.240–0.244; all paired bootstrap 95% CIs included zero).

**Conclusions:** Structured EHR data alone matched the discrimination and, after recalibration, the calibration of more complex multimodal fusion for 30-day COPD readmission prediction. These findings highlight the importance of rigorous temporal-leakage auditing in multimodal clinical prediction research and suggest that reformulating this task around temporal proximity to the outcome, rather than refining the fusion mechanism further, may be more productive for future work.

## Introduction

Chronic obstructive pulmonary disease (COPD) is a leading cause of morbidity and hospital resource utilization worldwide, and unplanned hospital readmission following a COPD-related admission remains a persistent challenge for both patient outcomes and health-system planning. A substantial body of work has sought to predict COPD exacerbation and readmission risk from routinely collected clinical data [1], with structured electronic health record (EHR) variables — laboratory values, vital signs, and diagnostic history — historically providing a consistent source of predictive signal.

A prevailing assumption in clinical artificial intelligence, sometimes termed the multimodal hypothesis, holds that combining structured data with additional modalities — imaging, physiological waveforms, or unstructured clinical narratives — should improve predictive performance by supplying diagnostic information inaccessible to any single modality alone [2]. This assumption has motivated a growing body of multimodal deep learning research in respiratory and critical care medicine, often without a corresponding emphasis on whether an additional modality’s contribution can be rigorously distinguished from noise, confounding, or data-leakage artifacts introduced during multimodal cohort construction.

Evidence on whether this assumption holds in practice is mixed. In a related study using a synthetic, Monte Carlo-generated COPD exacerbation cohort, we found that supplying raw, unstructured clinical notes directly to large language models, alongside structured physiological data, degraded rather than improved predictive performance relative to structured data alone — a phenomenon we termed the Clinical Notes Paradox [3]. That finding, however, was obtained in a specific representational regime: verbose, high-entropy free text supplied directly as model input, without intermediate compression or task-specific adaptation. Whether a comparable pattern holds when auxiliary modalities are instead represented as dense, frozen, pre-trained embeddings — a substantially different, and in much current multimodal clinical deep learning research more common, representational choice — has not, to our knowledge, been directly tested.

A second, largely separate concern in multimodal clinical prediction research using large public intensive-care and hospital datasets, such as the Medical Information Mart for Intensive Care (MIMIC) family of databases [4], is the risk of temporal data leakage: predictors drawn, however inadvertently, from a time window overlapping or following the outcome being predicted. Because such datasets aggregate years of longitudinal patient records across repeated hospital encounters for the same patients, constructing a leakage-free, admission-level multimodal cohort requires an explicit, verifiable temporal-alignment procedure — a step not always reported, or evidently performed, in prior multimodal work in this area.

The present study addresses both concerns for a specific, common clinical prediction task: 30-day, all-cause hospital readmission in patients with COPD. First, we conduct an explicit, empirically-verified audit of temporal data leakage across four modality-specific data-linkage pipelines (electrocardiogram, chest radiograph, clinical notes, and structured EHR data) derived from MIMIC-IV [4], [5]. Second, using frozen, pre-trained encoders for each modality — a dense-embedding representational regime distinct from the raw-text setting of the Clinical Notes Paradox study described above — we evaluate whether three independently-designed multimodal fusion architectures (naive concatenation, a scalar gated fusion mechanism, and an EHR-anchored residual fusion mechanism) improve 30-day readmission prediction over a frozen, structured-EHR-only baseline. Third, we assess not only discrimination but also probability calibration, a property of direct relevance to clinical risk-stratification use that is not consistently reported in comparable studies.

## Materials and Methods

This study is reported in accordance with the STROBE guideline for observational studies.

### Ethics statement

This study used data from the Medical Information Mart for Intensive Care IV (MIMIC-IV) database (version 3.1) [4], a publicly available, de-identified database of patients admitted to the Beth Israel Deaconess Medical Center (Boston, MA, USA). The original data collection and de-identification protocol for MIMIC-IV were approved by the Institutional Review Board of Beth Israel Deaconess Medical Center, which granted a waiver of informed consent given the retrospective and fully de-identified nature of the data. Because MIMIC-IV contains de-identified data, the investigator completed the required CITI “Data or Specimens Only Research” training and obtained database access permission through the PhysioNet Credentialed Health Data Use Agreement [6]; the certification record ID was 74100460. This study involved secondary analysis of publicly available, anonymized data, with no direct patient contact or access to identifiable private information; therefore, additional informed consent and ethics approval was not required.

Data for this study were accessed on 16 December 2025. The authors did not have access to information that could identify individual participants at any point during or after data collection, as MIMIC-IV is provided in a fully de-identified form prior to authorized access.

### Data source and cohort construction

MIMIC-IV is a large, publicly available electronic health record database derived from patients admitted to Beth Israel Deaconess Medical Center [4]. It contains de-identified demographic, diagnostic, laboratory, medication, and free-text clinical note data, in addition to linked chest radiograph imaging (MIMIC-CXR-JPG) and electrocardiogram waveform data (MIMIC-IV-ECG) for a subset of patients.

The patient cohort for this study was constructed using the MIMIC-IV-Data-Pipeline [5], configured for a non-ICU-index, 30-day hospital readmission prediction task, restricted to patients with a diagnosis of chronic obstructive pulmonary disease (COPD; ICD-10 code J44 and corresponding ICD-9 equivalents). From the resulting eligible population, a stratified sample of 2,000 patients was drawn, balanced by outcome (1,000 patients with a positive 30-day readmission outcome and 1,000 with a negative outcome; see “Outcome definition” below) to mitigate class imbalance during model training. Each patient contributed exactly one qualifying hospital admission, defined as the index admission from which the 30-day outcome window is measured; a one-to-one relationship between patients and index admissions was verified programmatically.

Of the 2,000 stratified patients, 1,279 (63.95%) had at least one linked hospital admission with usable structured electronic health record data (laboratory values, demographics, and clinical history) and were retained for the analytic cohort; the remaining 721 patients had a valid patient identifier in the source cohort file but no corresponding admission-level data (e.g., no linked hospital stay record) and were excluded prior to any modeling. The proportion of positive outcomes was similar among the excluded patients (about 346/721, 48%) and the retained patients (654/1,279, 51%).

The resulting 1,279-patient cohort was partitioned into training, validation, and test splits at the patient level (895 / 192 / 192 patients respectively), ensuring no patient appeared in more than one split.

The patient-selection process is summarized below:

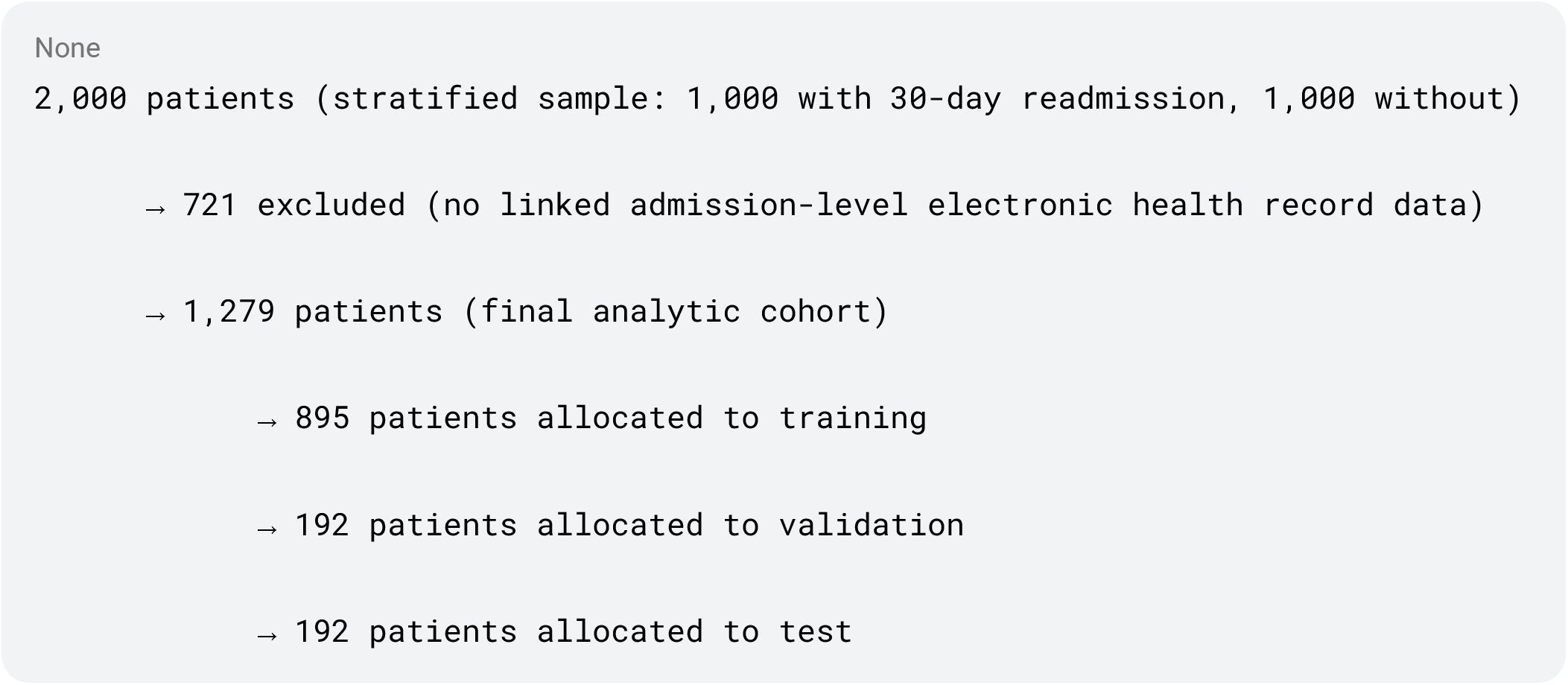

The stratified sample was drawn from the output of the MIMIC-IV-Data-Pipeline [5], whose intermediate selection outputs were not archived and could not be re-run; the natural prevalence of the outcome among eligible admissions was therefore estimated separately from the raw MIMIC-IV tables using an explicit, reproducible criterion: for each patient with at least one non-ICU hospital admission carrying a COPD diagnosis (ICD-9 491.x/492.x/496 or ICD-10 J44.x), the earliest such admission chronologically was treated as the index admission, and 30-day, all-cause readmission was determined identically to the definition in “Outcome definition” below. Under this criterion, 13,818 eligible non-ICU index admissions were identified, of which 2,506 (18.1%) met the 30-day readmission outcome — the natural, unbalanced prevalence in this population. The stratified 1,000/1,000 sample used in this study therefore substantially over-represents the positive class (50% versus an estimated 18.1% natural prevalence), a deliberate choice to support model training under class imbalance; this enrichment does not affect relative comparisons of discrimination between models evaluated on the same splits, but absolute risk estimates and calibration metrics, including Brier scores, are specific to the enriched cohort and should not be extrapolated to the unselected population.

### Outcome definition

The prediction target was 30-day, all-cause hospital readmission following discharge from the index admission, defined as a positive outcome if the patient had at least one subsequent hospital admission with an admission time occurring within 30 days after the discharge time of the index admission, and a negative outcome otherwise. This definition follows the standard “Readmission” task specification of the MIMIC-IV-Data-Pipeline [5] and is consistent with the broader hospital-readmission-prediction literature [7].

This outcome captures readmission for any cause and is not restricted to readmissions specifically attributable to a COPD exacerbation. A follow-up study using a validated administrative algorithm for identifying

COPD-exacerbation-specific readmissions is planned as a separate analysis (see Discussion).

### Temporal alignment and leakage prevention

Because the outcome is defined as an event occurring after discharge from the index admission (see “Outcome definition” above), any predictor derived from a given modality is valid only if it was observed at or before the discharge time (dischtime) of the index admission. Data recorded during or after a subsequent admission — including the readmission event that defines the outcome itself — constitutes temporal leakage and would artificially inflate predictive performance. This principle was applied uniformly across all four modalities: a modality-specific instance (an electrocardiogram recording, a chest radiograph, a clinical note, or a laboratory measurement) was retained as a predictor only if its associated timestamp fell within the closed interval [admittime, dischtime] of the patient’s verified index admission. Discharge summaries, which are authored at or shortly after discharge, were instead linked by exact index-admission identifier (see “Clinical notes”).

The index admission for each patient (subject identifier, hospital-admission identifier, admission time, and discharge time) was taken from the stratified cohort file described above, in which a one-to-one correspondence between patients and index admissions had been programmatically verified. This admission-window record served as the single source of truth against which every modality-specific linkage procedure was independently checked.

#### Leakage audit and corrections

Prior to finalizing the modality-specific linkage procedures described in the following section, each linkage procedure was audited by comparing the hospital-admission identifier and timestamp associated with every modality instance against the patient’s verified index-admission window. This audit identified three independent temporal-alignment issues, summarized in Table 1 and corrected before any model training or evaluation was performed.

**Table 1.**
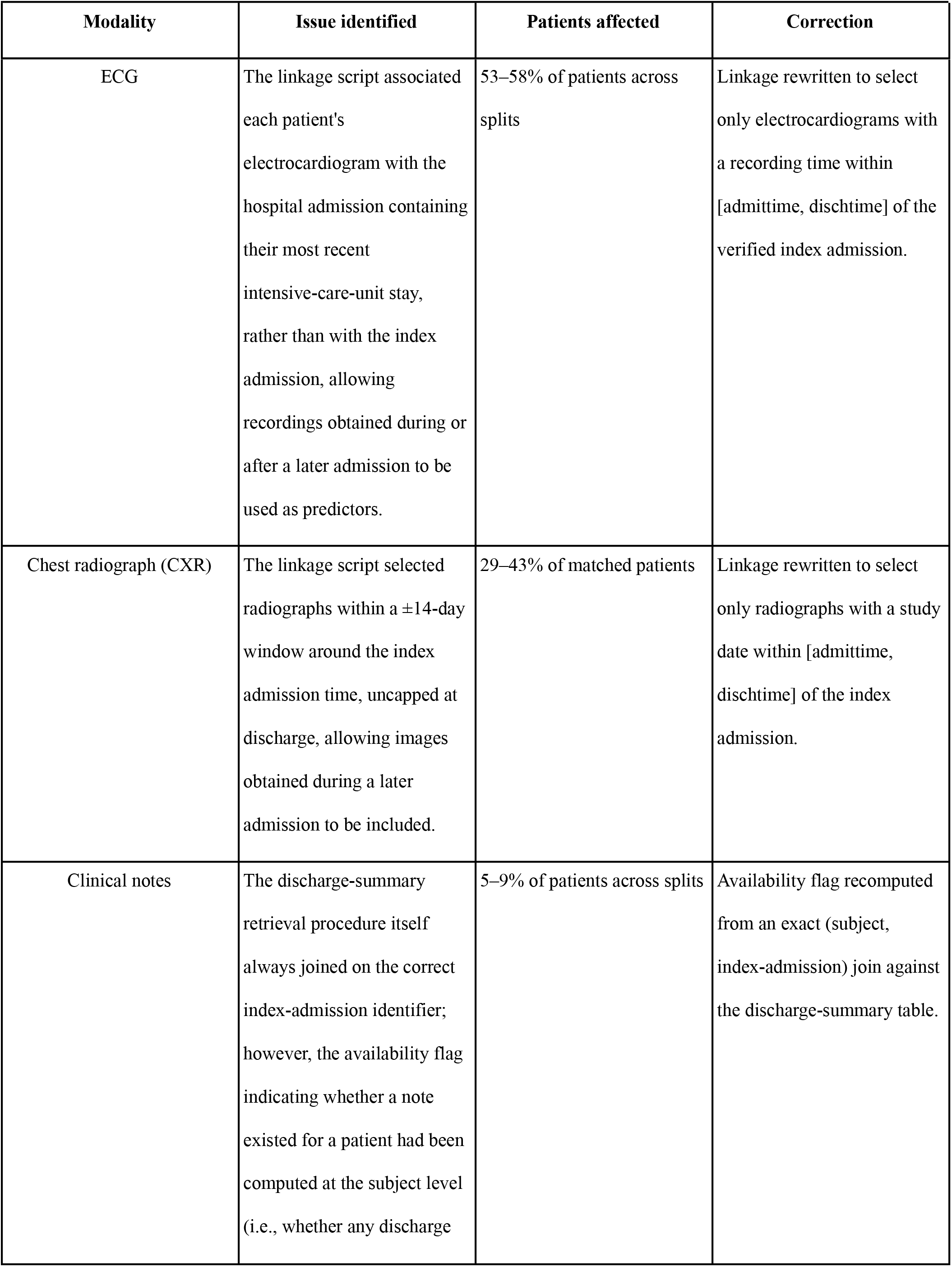
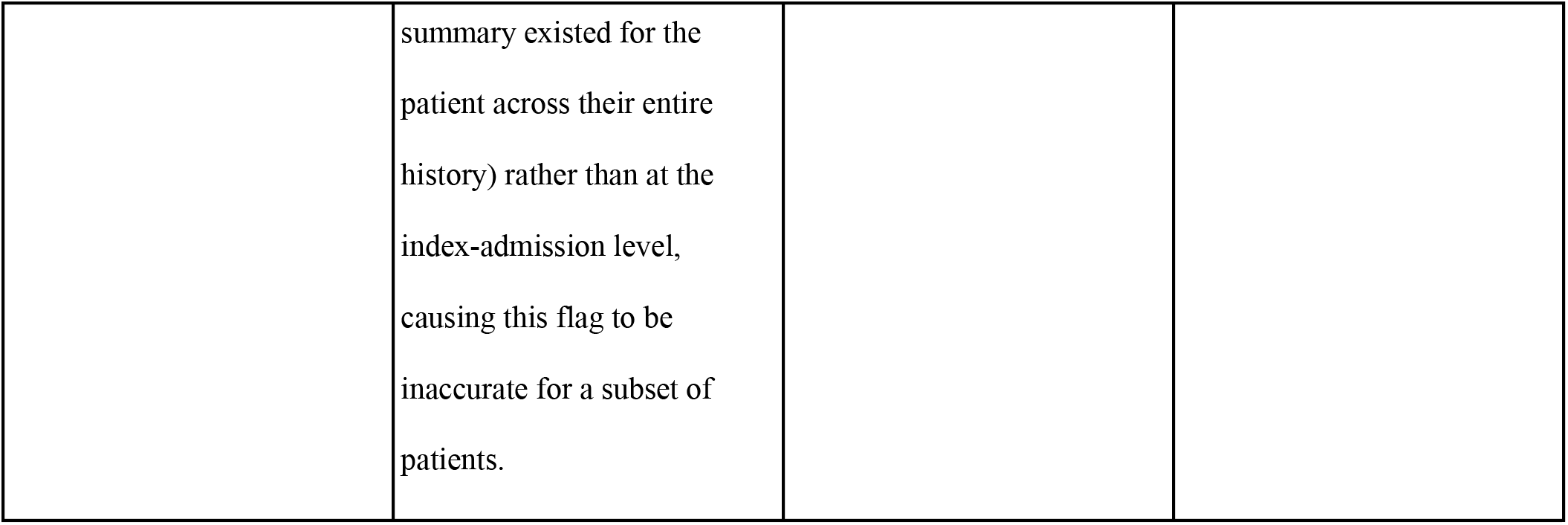
Temporal-leakage issues identified during the modality-linkage audit, patients affected, and corrections applied.

An analogous audit of the structured electronic-health-record (laboratory) data found only negligible temporal misalignment: 2 of 1,279 patients had a laboratory result recorded up to 2.4 hours after the index admission’s discharge time, consistent with a routine result-reporting delay for a specimen drawn during the admission rather than a genuine data leak; no correction was applied for this modality.

### Modality-specific preprocessing and frozen encoders

Four modality-specific encoders were used to generate patient-level feature representations from 12-lead electrocardiograms (ECG), free-text discharge summaries (clinical notes), chest radiographs (CXR), and structured electronic health record (EHR) data (laboratory values, demographics, and coded clinical history). Each encoder’s pre-trained backbone was kept entirely frozen throughout preprocessing and feature extraction, with no fine-tuning against the study outcome: this design choice was made to preserve consistency with each backbone’s self-supervised (or, for the tabular EHR sub-encoder, separately supervised — see “Electronic health record (EHR)” below) pretraining objective, and to limit the risk of overfitting given the small size of the analytic cohort (895 training patients) relative to the number of trainable parameters a full fine-tuning approach would introduce. Every modality-specific instance used as encoder input had already been restricted to the patient’s verified index-admission window (see “Temporal alignment and leakage prevention” above) prior to feature extraction.

#### Electrocardiogram (ECG)

Raw 12-lead ECG recordings from MIMIC-IV-ECG [8] contain linear redundancy across leads (Einthoven’s law, III = II − I; Goldberger’s augmented limb-lead equations for aVR, aVL, and aVF), so the underlying cardiac electrical field is fully captured by 8 independent leads (I, II, V1–V6). The preprocessing pipeline consisted of a 3rd-order Butterworth bandpass filter (0.5–45 Hz) to remove baseline wander and high-frequency electromyographic interference, channel-wise Z-score normalization, padding or cropping to a fixed 2,500-sample window, and polyphase resampling from the native 500 Hz to 250 Hz to match the input rate expected by the pre-trained encoder. The resulting input tensor had fixed dimensions of 8 × 2,500 (10 seconds of signal at 250 Hz).

Feature extraction used a frozen, pre-trained ECG-JEPA (Electrocardiogram Joint-Embedding Predictive Architecture) encoder [9], which maps the 8 × 2,500 input tensor to a native 768-dimensional embedding. No additional projection head was appended to the pre-trained encoder output, so as not to distort the pre-trained representation with randomly initialized, never-trained weights. An ECG was available within the verified index-admission window for 396/895 (44.2%) training, 83/192 (43.2%) validation, and 89/192 (46.4%) test patients; patients without a qualifying ECG were excluded from ECG-specific unimodal evaluation rather than assigned a zero-vector placeholder outside the fusion stage (see “Multimodal fusion architectures” below for how missing modalities are handled at fusion time).

#### Clinical notes

Discharge summaries were retrieved from MIMIC-IV-Note via an exact join on (subject identifier, index hospital-admission identifier), with no fallback to a note from a different admission if the exact match was unavailable (see “Temporal alignment and leakage prevention” above for the related admission-level availability-flag correction). MIMIC-IV-Note replaces protected health information with runs of underscore characters; because the target tokenizer split each such run into one sub-token per character, disproportionately consuming the encoder’s limited context window, runs of two or more underscores were collapsed to a single underscore prior to tokenization, preserving the redacted-content signal while reducing its token cost.

Discharge summaries were encoded with a frozen Bio_ClinicalBERT backbone [10], which has a 512-token input limit substantially shorter than the discharge summaries in this cohort. Following the general chunking approach used for hospital-readmission prediction from discharge summaries by Huang et al. [7], each note was split into overlapping chunks of up to 512 tokens (max_length=512, stride=128) using the tokenizer’s native sliding-window support; the 128-token stride follows a value reported explicitly for 512-token-limit clinical language models by Li et al. [11]. Each chunk was encoded independently, and the final hidden state of the [CLS] token was taken as the 768-dimensional chunk representation. Chunk representations were then pooled into a single note-level embedding; mean-pooling and max-pooling across chunks were both evaluated at the unimodal baseline stage (see “Unimodal baseline evaluation” below) and gave comparable near-chance performance, so mean-pooling was adopted as the representation carried forward into the multimodal fusion stage. A discharge summary was available within the index admission for 685/895 (76.5%) training, 141/192 (73.4%) validation, and 152/192 (79.2%) test patients.

#### Chest X-ray (CXR)

Chest radiographs were selected from MIMIC-CXR-JPG [12] using the corrected linkage described above (study date within [admittime, dischtime] of the verified index admission). A patient could have multiple valid radiographs within a single index admission.

Images were encoded with the frozen image encoder of BiomedCLIP [13] (512-dimensional native embedding). For patients with more than one valid radiograph, per-image embeddings were L2-normalized, mean-pooled across images, and L2-renormalized to yield a single patient-level embedding. Patients with no valid radiograph within the index admission received a zero-vector placeholder in the saved feature file, explicitly flagged via a separate boolean indicator and excluded — rather than treated as a real embedding — from any unimodal CXR evaluation. A valid radiograph was available for 136/895 (15.2%) training, 31/192 (16.1%) validation, and 25/192 (13.0%) test patients, the lowest coverage of the four modalities; this reflects the fact that chest radiography is a targeted diagnostic test ordered for a minority of admissions, not an artifact of the linkage procedure.

#### Electronic health record (EHR)

Unlike the other three modalities, the EHR representation combines two sub-encoders operating on the same per-admission record. A tabular branch encodes age, gender, ethnicity, and laboratory values (each paired with an explicit missingness-mask indicator) through a small multilayer perceptron (256 → 128 → 64 hidden units) into a 64-dimensional embedding (TabularMLP). A textual branch decodes each patient’s coded conditions, procedures, and medications from administrative codes into human-readable strings, serializes them into a single natural-language string per patient, and encodes this string with the same frozen Bio_ClinicalBERT backbone used for discharge summaries [10], using true token-level mean-pooling over the final hidden states rather than the [CLS]-token pooling used for discharge-summary chunks in “Clinical notes” above — a deliberate asymmetry between the two ClinicalBERT applications in this pipeline, noted here for transparency. The tabular (64-dimensional) and textual (768-dimensional) sub-embeddings are concatenated into a single 832-dimensional EHR representation. Because laboratory values, demographics, and coded clinical history are essentially always recorded for a hospital admission, EHR coverage is 100% across all three splits by design, in contrast to the substantially lower and clinically-driven coverage of ECG, notes, and CXR.

#### Methodological note on encoder asymmetry

Unlike the ECG, clinical-notes, and CXR encoders, which are frozen, purely self-supervised backbones that never had access to the study outcome, the tabular sub-encoder (TabularMLP) was itself trained in a supervised fashion against the 30-day readmission target — with training, validation, and test splits respected throughout — before being frozen and used solely for embedding extraction, functionally similar to a linear probe with one additional trainable hidden layer. This does not constitute leakage, but it is a structural asymmetry across the four modality encoders that is disclosed explicitly here, since it may partly explain the EHR modality’s comparatively strong unimodal performance reported in “Unimodal baseline evaluation” below.

### Unimodal baseline evaluation

Before evaluating any multimodal fusion architecture, the predictive signal available from each modality in isolation was assessed with a frozen linear probe: an L2-regularized logistic regression classifier (inverse regularization strength C=1.0, class_weight=’balanced’, random_state=42, max_iter=1000) trained directly on the native, frozen embedding for that modality. Features were standardized (zero mean, unit variance) using statistics computed on the training split only, then applied unchanged to the validation and test splits. This single, fixed configuration — with no hyperparameter tuning — was applied identically across all four modalities, so that no modality’s baseline benefited from configuration choices unavailable to the others.

For the three modalities with incomplete coverage (ECG, clinical notes, CXR), only patients with a valid, leakage-audited embedding for that specific modality were included in the corresponding modality-specific training, validation, and test sets (see Table 2 for exact counts); patients without a qualifying instance were excluded from that modality’s unimodal evaluation rather than represented with a zero-vector placeholder, since a placeholder value carries no genuine modality-specific information and would bias a linear probe’s coefficients toward detecting missingness itself rather than genuine signal. The EHR modality, with 100% coverage by design, required no such exclusion.

**Table 2.** Frozen backbone, native embedding dimensionality, and per-split coverage for each of the four modality-specific encoders.

| Modality | Backbone | Embedding dim. | Train coverage | Val. coverage | Test coverage |
| --- | --- | --- | --- | --- | --- |
| ECG | ECG-JEPA [9] | 768 | 396/895 (44.2%) | 83/192 (43.2%) | 89/192 (46.4%) |
| Clinical notes | Bio_ClinicalBERT [10] | 768 | 685/895 (76.5%) | 141/192 (73.4%) | 152/192 (79.2%) |
| CXR | BiomedCLIP [13] | 512 | 136/895 (15.2%) | 31/192 (16.1%) | 25/192 (13.0%) |
| EHR (tabular + text) | TabularMLP + Bio_ClinicalBERT [10] | 64 + 768 = 832 | 895/895 (100%) | 192/192 (100%) | 192/192 (100%) |

#### Non-linear adapter probes

Because a linear probe can only reveal a signal that is linearly separable in the frozen embedding space, a near-chance linear-probe result does not, by itself, establish that a modality carries no exploitable signal for the outcome — it may instead reflect a limitation of the linear probe itself. To address this, a lightweight, parameter-efficient non-linear adapter [14] was additionally trained and evaluated for a subset of modalities. The adapter consisted of a bottleneck transformation (a down-projection, a GELU nonlinearity, dropout, and an up-projection back to the native embedding dimensionality) added residually to the frozen embedding, with the up-projection initialized near zero so that the adapter began training as a near-identity transformation of the frozen embedding. The adapter and a linear classification head were trained jointly, using the same class-balanced loss as elsewhere in this study, with a substantially larger number of full-batch optimization steps than the standard training protocol described below, to ensure convergence; convergence was confirmed by inspecting the training-loss trajectory for each run.

This adapter probe was applied to the ECG and clinical-notes modalities. An equivalent attempt for the CXR modality was not considered reliable: given the small number of available training patients for this modality (136; see Table 2), the adapter’s training loss collapsed to near zero, indicating substantial memorization of the training set, and the resulting test-set estimate (based on only 25 patients) was judged too unstable to interpret as evidence either for or against genuine signal in the CXR modality. This result is reported in the Results section as inconclusive due to sample size, and is not treated as a negative finding about the CXR modality’s content.

### Multimodal fusion architectures

Three multimodal fusion architectures were compared, each combining the four frozen modality embeddings described above (Table 2) into a single patient-level prediction. All three architectures share an identical per-modality projection component: a learned linear layer, followed by layer normalization, a ReLU nonlinearity, and dropout, mapping each modality’s native embedding to a common 32-dimensional shared space (see “Training protocol and hyperparameter optimization” below for how this dimensionality was selected). Holding this projection component identical across all three architectures, and using a single linear layer as the final classification head in every case, was a deliberate design choice: it isolates the fusion mechanism itself — rather than differences in projection or classifier capacity — as the only architectural variable being compared.

#### Naive concatenation

The first architecture concatenates the four projected, 32-dimensional modality embeddings into a single 128-dimensional vector, which is passed directly to the linear classification head. This architecture has no explicit mechanism for handling missing modalities: a patient missing a given modality is represented by a zero vector in the corresponding segment of the concatenation, and the model must learn, implicitly and without any explicit signal indicating which segments are missing, to discount these segments appropriately. This architecture serves as the reference point for evaluating whether the more explicit missingness-handling mechanisms described below provide a measurable advantage over simple concatenation.

#### Scalar gated fusion

The second architecture computes a learned scalar weight for each modality, for each patient, and combines the projected embeddings as a weighted sum rather than a concatenation. This follows the general design of Gated Multimodal Units [15], generalized here from the original two-modality formulation to four modalities. For each modality, a linear layer maps its projected embedding to a single scalar gate score; the four scores are then combined with a softmax function to yield normalized, non-negative weights summing to one, and the fused representation is the weighted sum of the four projected embeddings using these weights.

#### Handling of missing modalities

A key design choice distinguishes this architecture’s handling of missing modalities from the naive concatenation approach above: for a given patient, any modality that is missing is explicitly excluded from the softmax computation itself (by setting its gate score to negative infinity before the softmax is applied), rather than being included with a zero-vector embedding and left for the softmax to implicitly down-weight. This guarantees that a missing modality’s gate weight is exactly zero by construction, and that its (arbitrary, placeholder) embedding value can never influence the fused representation. This design was adopted specifically to avoid a failure mode identified earlier in this study (see “Temporal alignment and leakage prevention” above): if a model is allowed to observe a zero-vector placeholder without an explicit indicator that the corresponding modality is absent, it risks learning to treat the mere presence of a zero-vector as a predictive feature in itself — a spurious correlation with missingness, rather than with genuine modality content — the same category of risk that had been found, in a different form, to affect the original (pre-correction) ECG linkage in this study.

In addition to the fused representation and the classification output, this architecture returns the per-patient, per-modality gate weights themselves, which were used for a post hoc interpretability analysis (see Results).

#### EHR-anchored residual fusion

The third architecture was designed specifically in response to an empirical finding from the unimodal baseline evaluation (see Results): that the EHR modality alone substantially outperformed every other individual modality, and that the two fusion architectures described above did not reliably exceed the EHR-alone baseline’s performance. This architecture treats the EHR modality — always available for 100% of patients (Table 2) — as a fixed representational anchor, and the other three modalities as a source of learned correction applied on top of this anchor, following the general principle of residual learning [16]: the auxiliary correction can be learned down to exactly zero, at which point the architecture’s output reduces exactly to a function of the EHR embedding alone, rather than merely approximating it.

Concretely, the projected EHR embedding serves as the anchor representation. The projected embeddings of the other three modalities are combined into a single auxiliary representation using the same masked-softmax mechanism described above for scalar gated fusion, restricted to these three modalities only (the EHR modality does not compete in this softmax, since it is the anchor rather than a candidate to be weighted against the others). A single learned scalar gate — a sigmoid-activated function of both the anchor and the auxiliary representation — then determines how much of the auxiliary representation is added to the anchor to form the final fused representation. For patients with none of the three auxiliary modalities available, the auxiliary representation is exactly the zero vector by construction, and the fused representation reduces exactly to the anchor — a structural guarantee, rather than a learned approximation, that such patients’ predictions depend only on their EHR data.

### Training protocol and hyperparameter optimization

#### Common training protocol

All three fusion architectures were trained with the same cross-entropy loss, with class weights reproducing the standard formula weight[c] = n_samples / (n_classes × n_samples_c) — the same class-balancing approach used for the unimodal logistic-regression baselines above — and no additional per-patient weighting scheme (e.g., no up-weighting of patients with more modalities available), to avoid biasing training toward the minority subgroup of patients with near-complete modality coverage. Models were optimized with Adam, with early stopping on validation-set ROC-AUC (patience of 20 epochs without improvement, maximum 200 epochs). Every architecture was trained and evaluated across five random seeds (0–4); for formal statistical comparisons (see “Statistical analysis” below), test-set predictions were computed as an ensemble, averaging the predicted probability across the five corresponding seed-specific model checkpoints.

#### Hyperparameter search

A two-stage grid search was conducted for the naive concatenation and scalar gated fusion architectures, applying an identical search space and protocol to both to preserve a fair comparison. In the first stage, the shared projection dimensionality (32, 64, or 128), learning rate (1e−4, 3e−4, or 1e−3), and weight decay (1e−4, 1e−3, or 1e−2) were searched jointly (27 configurations), each evaluated across three seeds. In the second stage, with the first stage’s best configuration held fixed, the projector’s dropout probability (0.1, 0.2, or 0.3) and training batch size (8, 16, or 32) were searched jointly (9 configurations), again each evaluated across three seeds. (Full per-seed results for every individually-searched configuration in both stages are provided in S2 Table.)

#### Selection criterion

Configurations were ranked by the mean validation ROC-AUC across the three search seeds minus its standard deviation, rather than by the mean alone. This criterion was adopted after an initial mean-only selection, for the naive concatenation architecture, favored a configuration that scored well across the three search seeds but proved unstable on full five-seed confirmation and independent test-set evaluation (see “Statistical analysis” below for the specific instability observed and how it was diagnosed); selecting instead by mean-minus-standard-deviation — a simple form of lower-confidence-bound selection that penalizes variance directly — favors configurations that are reliably good across random initializations rather than merely fortunate on a small number of search seeds, and was applied retrospectively to re-select both architectures’ final configurations.

The resulting configurations were: shared projection dimensionality of 32 for both architectures; for naive concatenation, a learning rate of 1e−4, weight decay of 1e−2, dropout of 0.2, and batch size of 16; for scalar gated fusion, a learning rate of 1e−3, weight decay of 1e−3, dropout of 0.1, and batch size of 32. Each final configuration was then confirmed with the full five-seed training and evaluation protocol described above.

The EHR-anchored residual fusion architecture was not independently searched over this space, given the time constraints of the present study; it reused naive concatenation’s selected configuration as a reasonable starting point. This is disclosed here as a limitation specific to the three-way architectural comparison: unlike the naive-versus-gated comparison, which benefited from an identical and independently-optimized configuration for both architectures, the residual fusion architecture’s results should be interpreted with the possibility in mind that its own optimal configuration may differ from the one used.

### Statistical analysis

#### Primary and secondary metrics

Discrimination was summarized using the area under the receiver operating characteristic curve (ROC-AUC) and the area under the precision-recall curve (PR-AUC), both of which are independent of any specific classification threshold and were treated as the primary metrics throughout this study. Accuracy, sensitivity, specificity, and F1-score were also computed, using a fixed decision threshold of 0.5 on the predicted probability with no post hoc recalibration; because these metrics depend on this specific, uncalibrated threshold, they are reported as secondary metrics only, alongside the threshold-independent primary metrics, and are not used as the basis for any comparative claim between models in this study.

#### Significance testing

Formal comparisons between models were conducted with a paired, patient-level bootstrap procedure on the test set, using each model’s five-seed ensemble prediction (the mean predicted probability across the five seed-specific checkpoints described above). For a given pair of models, 10,000 bootstrap resamples of the test-set patients were drawn with replacement (using the same resampled indices for both models in a pair, to preserve pairing); for each resample, the difference in ROC-AUC between the two models was computed, and any resample in which the sampled labels contained only one class was discarded. The resulting distribution of differences was used to construct a 95% confidence interval (2.5th and 97.5th percentiles) and a two-sided p-value proxy (twice the smaller of the proportion of resamples favoring each model). This procedure was applied to two sets of comparisons: naive concatenation versus scalar gated fusion, and the EHR-alone baseline versus each of the three fusion architectures in turn.

An unexpected result during model-selection confirmation (a single random seed of the naive concatenation architecture showing a sensitivity and F1-score of zero on the test set; see Results) was investigated before being accepted as a genuine finding: that seed’s ROC-AUC (a threshold-independent measure) remained above chance, indicating that the apparent collapse in the secondary, threshold-dependent metrics reflected a shift in the fixed 0.5 decision boundary’s suitability for that particular seed’s predicted-probability distribution, rather than a genuine loss of discriminative ability. This observation motivated the primary/secondary metric distinction described above and is reported in the Results as a methodological note rather than a substantive finding about the naive concatenation architecture.

#### Missingness-robustness stress test

To test whether the scalar gated fusion architecture’s explicit missing-modality handling translates into greater robustness under missingness more severe than the dataset’s naturally-occurring pattern, both the naive concatenation and scalar gated fusion architectures (five-seed ensembles) were re-evaluated on the same test-set patients under four cumulative levels of artificially-imposed modality removal: the natural missingness pattern; CXR forced unavailable for every patient; CXR and ECG forced unavailable; and CXR, ECG, and clinical notes forced unavailable (EHR-only). At each level, the embeddings and availability masks for the removed modalities were set to zero and false, respectively, for every patient, regardless of whether that modality was naturally available — ensuring the manipulation was applied identically and correctly to both architectures irrespective of whether a given architecture makes explicit use of the availability mask.

#### Calibration analysis

In addition to discrimination, model calibration — the agreement between predicted probabilities and observed outcome frequencies — was assessed for the EHR-alone baseline and all three fusion architectures on the test set, using the Brier score (the mean squared difference between predicted probability and observed binary outcome, for which lower values indicate better calibration) and reliability diagrams. Because the test set contains only 192 patients, reliability diagrams were constructed using quantile-based binning (equal numbers of patients per bin) rather than fixed-width probability bins, to avoid bins with very few or zero patients. Because the Brier score combines discrimination and calibration, we additionally reported the calibration slope (the coefficient of a logistic regression of the outcome on the logit of the predicted probability, fitted on the test set; 1 indicates ideal calibration) and the Brier skill score relative to a constant predictor at the test prevalence. To test whether any difference in calibration persisted after simple post-hoc recalibration, a two-parameter logistic recalibration (outcome regressed on the logit of the predicted probability) was fitted on the validation set for each model (fusion models as five-seed ensembles, EHR-alone as a single model) and applied unchanged to the test set. The difference in Brier score between EHR-alone and each fusion architecture was then compared with a paired patient-level bootstrap (10,000 resamples, 95% percentile interval).

### Software and code availability

All preprocessing, feature extraction, model training, and analysis code was implemented in Python 3.12.13, using PyTorch 2.11.0 and torchvision 0.26.0 for model implementation and training, the Hugging Face transformers library (version 5.12.1) for the Bio_ClinicalBERT backbone, open_clip_torch (version 3.3.0) for the BiomedCLIP backbone, and scikit-learn (version 1.8.0) for the unimodal logistic-regression baselines, feature standardization, calibration analysis, and evaluation metrics. Additional dependencies included numpy (2.4.3), pandas (3.0.1), scipy (1.17.1, used for ECG bandpass filtering and resampling), wfdb (4.3.1, used for reading raw ECG waveform records), Pillow (12.1.1, used for chest radiograph loading), matplotlib (3.11.0, used for the calibration reliability diagram), and tqdm (4.67.3). All experiments were run on macOS (arm64) using PyTorch’s Metal Performance Shaders (MPS) backend for GPU acceleration.

Code implementing the cohort-enrichment and temporal-leakage-correction scripts, the four modality-specific encoders, the three fusion architectures, the training and hyperparameter-search procedures, and all statistical analyses described above is available at https://doi.org/10.5281/zenodo.23096348. As described in the Data Availability Statement, the underlying MIMIC-IV data cannot be redistributed directly; the repository allows other credentialed PhysioNet users to reproduce the cohort extraction and full analysis pipeline described in this manuscript.

### Data and code availability

MIMIC-IV, MIMIC-IV-Note, MIMIC-CXR-JPG and MIMIC-IV-ECG are available to credentialed users through PhysioNet and cannot be redistributed by the authors. The analysis code is deposited on Zenodo (https://doi.org/10.5281/zenodo.23096348) with restricted access, which is granted on request to credentialed PhysioNet users.

## Results

Across every analysis reported below, the same held-out test set (192 patients, unseen by any training or model-selection decision) was used, and results are reported for the four unimodal baselines, the EHR-alone reference model, and the three multimodal fusion architectures described in Materials and Methods.

### Cohort characteristics

Table 3 summarizes the demographic and outcome characteristics of the 1,279-patient analytic cohort, overall and by split. The median age was approximately 70 years, and the cohort was balanced by gender (52.7% male, 47.3% female). Consistent with the demographic composition of the source institution, the majority of patients were recorded as White (78.8%), with smaller proportions recorded as Black/African American (9.2%), Hispanic/Latino (2.7%), Asian (0.9%), and other or unknown race (8.6%). The 30-day readmission outcome was reasonably balanced across splits (51.2% in training, 47.4% in validation, 54.7% in test), consistent with the stratified 1,000/1,000 sampling design described in “Data source and cohort construction.”

**Table 3.**
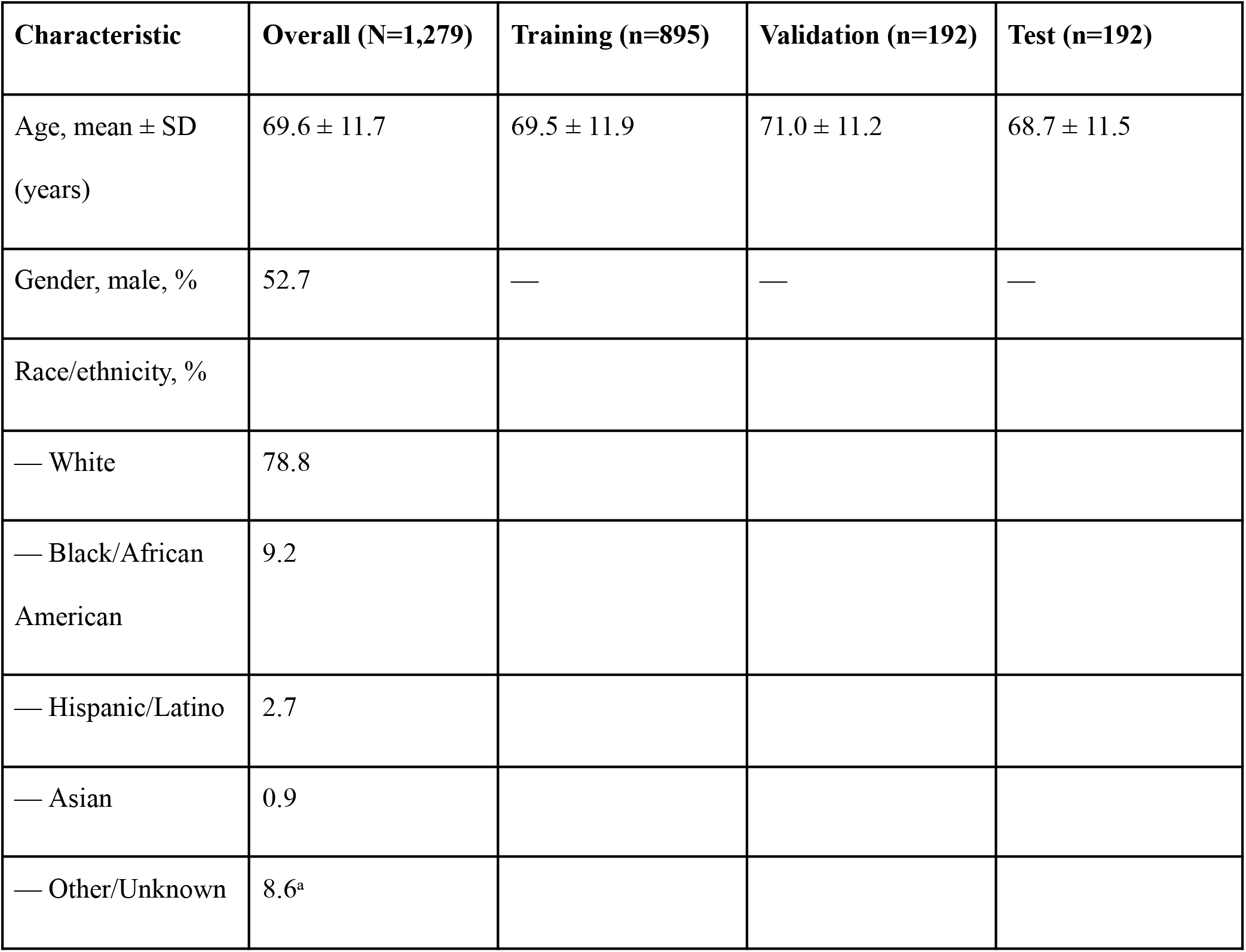
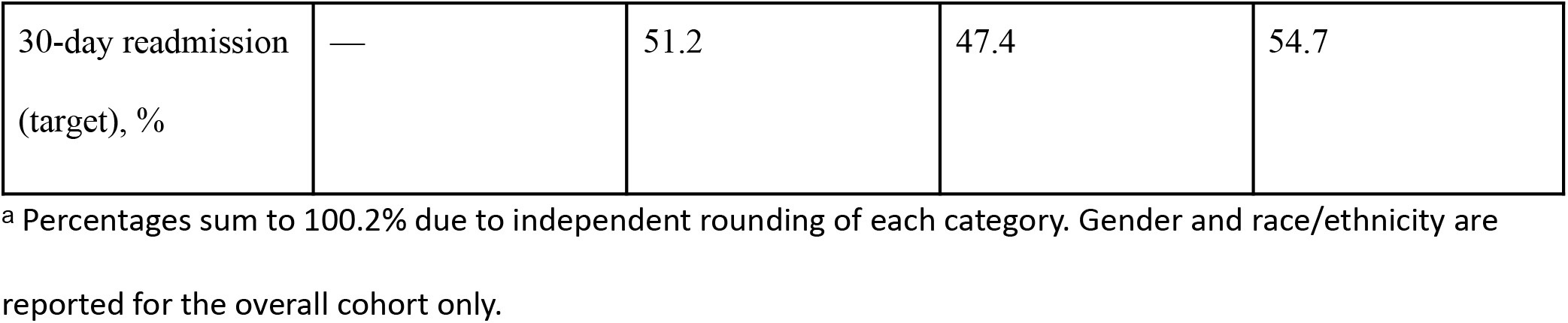
Demographic and outcome characteristics of the analytic cohort.

### Unimodal baseline evaluation

Table 4 reports the test-set discrimination of the frozen linear probe for each modality in isolation. Every modality except EHR performed close to chance level (ROC-AUC near 0.50); the EHR-alone probe substantially outperformed the other three.

**Table 4.** Unimodal frozen linear-probe discrimination on the test set.

| Modality | N (test) | ROC-AUC | PR-AUC |
| --- | --- | --- | --- |
| ECG | 89 | 0.4802 | — |
| Clinical notes (mean-pooled) | 152 | 0.4957–0.5307<sup>a</sup> | — |
| CXR | 56 (val+test pooled)<sup>b</sup> | 0.4628 | — |
| EHR (tabular + text) | 192 | 0.6343 | 0.6677 |

<sup>a</sup> Range reflects mean-pooling versus max-pooling across note chunks (see Materials and Methods); both gave comparable near-chance performance.
<sup>b</sup> CXR is reported on the pooled validation+test set (N=56) rather than test alone (N=25), given the modality's low coverage; see Materials and Methods for the corresponding caveat on statistical power.

### Non-linear adapter probes

To rule out insufficient linear-probe capacity as an explanation for the near-chance unimodal results above, a non-linear bottleneck adapter was additionally evaluated for the ECG and clinical-notes modalities (Table 5). The ECG adapter result closely reproduced the linear-probe finding, despite the adapter’s training loss reaching near-total convergence, ruling out under-fitting as an explanation. The clinical-notes adapter showed a modest improvement over the linear probe. The CXR adapter result is reported for completeness but is not interpreted as evidence for or against genuine signal in that modality: with only 136 training and 25 test patients, its training loss collapsed to near zero (0.006), indicating substantial memorization, and the resulting test-set estimate is considered statistically unreliable.

**Table 5.**
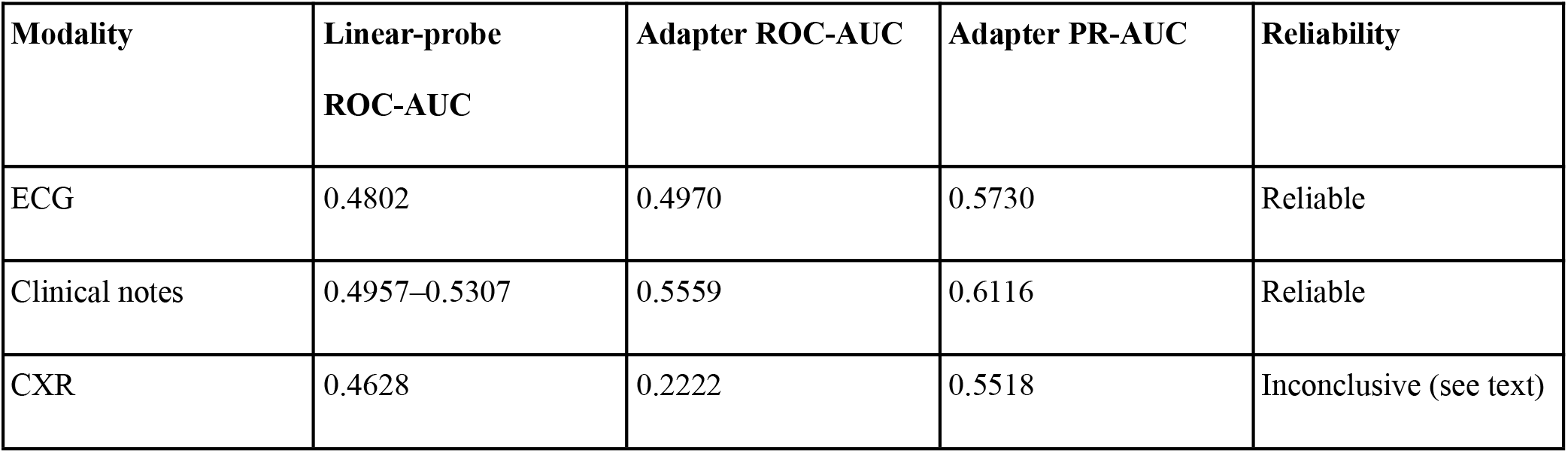
Non-linear adapter probe results, compared to the corresponding linear-probe baseline.

| Modality | Linear-probe ROC-AUC | Adapter ROC-AUC | Adapter PR-AUC | Reliability |
| --- | --- | --- | --- | --- |
| ECG | 0.4802 | 0.4970 | 0.5730 | Reliable |
| Clinical notes | 0.4957–0.5307 | 0.5559 | 0.6116 | Reliable |
| CXR | 0.4628 | 0.2222 | 0.5518 | Inconclusive (see text) |

### Multimodal fusion versus the EHR-alone baseline

Table 6 reports test-set discrimination for the EHR-alone baseline and each of the three fusion architectures, both as the mean ± standard deviation across five independently-evaluated seeds and, for the formal statistical comparisons below, as the discrimination of each architecture’s five-seed ensemble prediction.

**Table 6.** Test-set discrimination for the EHR-alone baseline and the three fusion architectures. (Full per-seed results for all three fusion architectures are provided in S1 Table.)

| Model | Mean ± SD ROC-AUC (5 seeds) | Mean ± SD PR-AUC (5 seeds) | 5-seed ensemble ROC-AUC |
| --- | --- | --- | --- |
| EHR-alone | — (single model) | — (single model) | 0.6343 |
| Naive concatenation | $0.6031 \pm 0.0238$ | $0.6233 \pm 0.0177$ | 0.6228 |
| Scalar gated fusion | $0.5925 \pm 0.0126$ | $0.6338 \pm 0.0136$ | 0.5956 |
| EHR-anchored residual fusion | $0.5853 \pm 0.0363$ | $0.6373 \pm 0.0469$ | 0.6028 |

#### Significance testing

Paired bootstrap testing (10,000 resamples) found no statistically significant difference between naive concatenation and scalar gated fusion (mean difference −0.0270, 95% CI [−0.0731, +0.0185], p = 0.2498), nor between the EHR-alone baseline and any of the three fusion architectures (EHR-alone minus scalar gated fusion: +0.0391, 95% CI [−0.0459, +0.1256], p = 0.374; EHR-alone minus naive concatenation: +0.0121, 95% CI [−0.0694, +0.0936], p = 0.770; EHR-alone minus residual fusion: +0.0319, 95% CI [−0.0546, +0.1186], p = 0.466). Every confidence interval included zero. Notably, the point estimate favored the EHR-alone baseline in all three comparisons against a fusion architecture, a consistent pattern across three independently-designed fusion mechanisms.

### Missingness-robustness stress test

Table 7 reports the five-seed ensemble ROC-AUC for naive concatenation and scalar gated fusion under increasing, artificially-imposed modality removal. Contrary to the expectation that the explicit missing-modality handling of scalar gated fusion would degrade more gracefully than naive concatenation’s implicit handling, both architectures’ discrimination improved as auxiliary modalities were removed, with the EHR-only condition the best-performing condition for both architectures; the gap between the two architectures widened, in naive concatenation’s favor, as missingness increased.

**Table 7.** Test-set ROC-AUC under increasing artificially-imposed modality removal.

| Severity level | Scalar gated fusion | Naive concatenation | Gap (gated – naive) |
| --- | --- | --- | --- |
| Natural missingness (all modalities as available) | 0.5956 | 0.6228 | −0.0271 |
| CXR forced unavailable | 0.5999 | 0.6375 | −0.0377 |
| CXR + ECG forced unavailable | 0.5999 | 0.6407 | −0.0408 |
| CXR + ECG + notes forced unavailable (EHR-only) | 0.6010 | 0.6476 | −0.0466 |

### Calibration

Fig 2 shows the test-set reliability diagram for the EHR-alone baseline and the three fusion architectures, before and after recalibration. The raw EHR-alone probabilities were the least well calibrated (Brier score 0.3203; calibration slope 0.12), with predicted probabilities systematically more extreme than the observed event rates, whereas the fusion architectures had Brier scores of 0.241–0.246 (calibration slopes 0.60–0.91) (Table 8).

**Fig 1.**
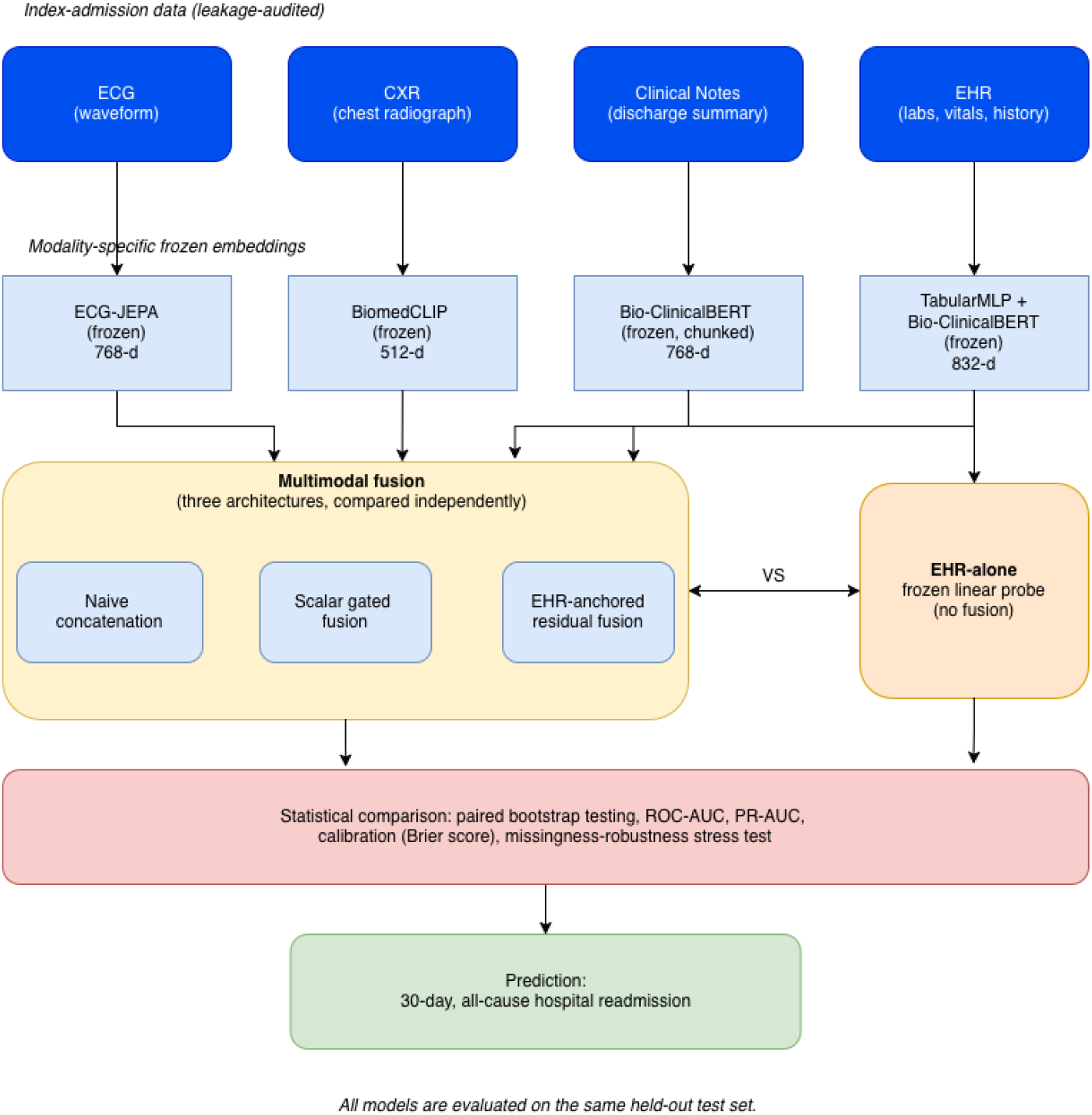
Schematic overview of the study pipeline. Four input modalities from the patient’s index admission are encoded by frozen, pre-trained, modality-specific encoders. Three multimodal fusion architectures (naive concatenation, scalar gated fusion, and EHR-anchored residual fusion) are evaluated under an identical protocol and, together with a reference EHR-alone frozen linear probe, compared via paired bootstrap significance testing, calibration analysis, and a missingness-robustness stress test. All models are evaluated on the same held-out test set.

**Fig 2.**
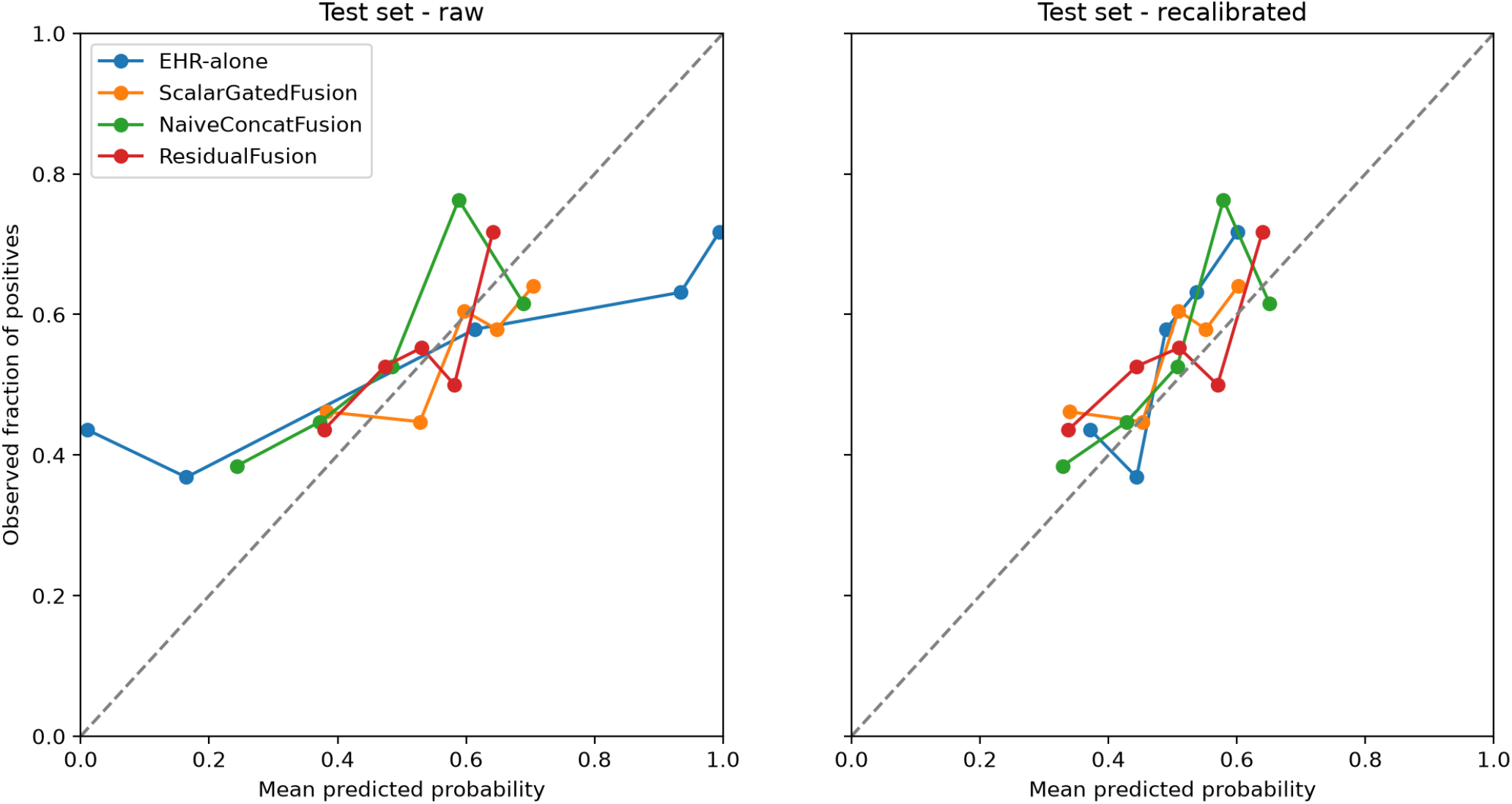
Calibration (reliability) diagrams on the test set for the EHR-alone baseline and the three fusion architectures, using quantile-based binning: raw predicted probabilities (left) and after logistic recalibration fitted on the validation set (right). The dashed diagonal indicates perfect calibration.

**Table 8.** Brier score and calibration of the EHR-alone baseline and the three fusion architectures on the test set, before and after logistic recalibration fitted on the validation set. Brier skill = 1 − Brier / [p(1−p)], where the reference is a constant predictor at the test prevalence (p = 0.547; Brier 0.2478). Fusion rows are 5-seed ensembles; EHR-alone is a single model.

| Model | Raw Brier | Raw calibration slope | Recalibrated Brier | Recalibrated Brier skill |
| --- | --- | --- | --- | --- |
| EHR-alone | 0.3203 | 0.12 | 0.2388 | 0.036 |
| Scalar gated fusion | 0.2427 | 0.69 | 0.2439 | 0.016 |
| Naive concatenation | 0.2455 | 0.60 | 0.2398 | 0.032 |
| EHR-anchored residual fusion | 0.2411 | 0.91 | 0.2430 | 0.019 |

Because the Brier score combines discrimination and calibration, and because the test prevalence (54.7%) implies a Brier score of 0.248 for a constant predictor, we examined whether this difference persisted after post-hoc recalibration. A two-parameter logistic recalibration (outcome regressed on the logit of the predicted probability) was fitted on the validation set for each model and applied unchanged to the test set. After recalibration, the Brier score of EHR-alone fell to 0.2388, comparable to that of the fusion models (0.2398–0.2439); paired bootstrap differences included zero in all three comparisons (EHR-alone minus scalar gated fusion −0.0052, 95% CI [−0.0183, +0.0080]; minus naive concatenation −0.0011, [−0.0160, +0.0138]; minus residual fusion −0.0043, [−0.0191, +0.0105]). All models remained only marginally better than a constant predictor (Brier skill 0.016–0.036). The raw calibration gap therefore reflected overconfidence of the EHR-alone probe rather than an advantage of multimodal fusion.

### Gate-weight interpretability

The scalar gated fusion architecture’s learned per-modality gate weights were inspected on the test set, aggregated across the five seed-specific models (Table 9). The gate assigned the EHR modality a substantially higher weight than the other three modalities on average, and specifically among the 22 test patients with all four modalities available — the pattern in which the gate faces a genuine four-way choice — the EHR modality alone received more weight than the other three modalities combined. This pattern was learned entirely from the classification loss, with no explicit supervision indicating which modality should matter, and is directly consistent with the unimodal baseline and fusion-comparison results reported above.

**Table 9.** Mean scalar gate weight per modality on the test set, overall and by modality-availability pattern.

| Availability pattern | ECG | Notes | CXR | EHR |
| --- | --- | --- | --- | --- |

| (N patients) |  |  |  |  |
| --- | --- | --- | --- | --- |
| Overall mean weight when present | 0.1763 | 0.1826 | 0.1921 | 0.7487 |
| EHR only (N=40) | — | — | — | 1.0000 |
| Notes + EHR (N=60) | — | 0.211 | — | 0.789 |
| ECG + Notes + EHR (N=67) | 0.186 | 0.172 | — | 0.642 |
| All 4 modalities (N=22) | 0.146 | 0.140 | 0.188 | 0.527 |

## Discussion

### Principal findings

This study evaluated whether combining ECG, chest radiograph, and clinical-note data with structured electronic health record (EHR) data, via three distinct multimodal fusion architectures, improves prediction of 30-day hospital readmission in COPD patients beyond what EHR data alone achieves. Three findings emerged. First, an independent, empirical audit of the four modality-linkage pipelines identified and corrected three previously undetected temporal-leakage issues (Table 1), affecting between 5% and 58% of patients depending on the modality. Second, once corrected, every modality except EHR carried a near-chance discriminative signal in isolation (Table 4), a finding that a non-linear adapter probe did not overturn for ECG and only modestly qualified for clinical notes (Table 5). Third, none of the three independently-designed multimodal fusion architectures achieved a statistically significant improvement over the EHR-alone baseline (Table 6), and a missingness-robustness stress test found that removing the auxiliary modalities improved, rather than degraded, both tested architectures’ discrimination (Table 7) — the auxiliary modalities appear to be, at best, neutral and at worst mildly detrimental contributors in this setting. Fourth, the lower raw Brier scores of the fusion architectures did not survive recalibration and reflected overconfidence of the EHR-alone probe rather than a benefit of fusion.

### Interpretation

A plausible explanation for this pattern relates to the temporal proximity of each modality’s information content to the prediction target. EHR laboratory values and vital signs recorded close to discharge directly reflect a patient’s physiological state at the point a readmission-risk assessment would be made in practice. By contrast, the ECG, chest radiograph, and clinical note available for a given patient reflect a single snapshot drawn from within the index admission, which may occur substantially earlier than discharge and need not be temporally or clinically proximal to whatever specific event precipitates a readmission within the following 30 days, since the outcome definition used here is not restricted to readmissions of any particular cause (see “Outcome definition” and the corresponding limitation below). This interpretation is consistent with the broader hospital-readmission-prediction literature [7], in which structured clinical variables measured close to discharge are established strong predictors of readmission risk.

### Comparison with prior literature

The magnitude of the EHR-alone baseline’s discrimination (Table 4) is broadly consistent with the modest-to-moderate effect sizes typically reported for readmission-prediction models built on structured data alone. We do not attempt a precise numerical comparison to any single prior study’s reported metrics, since differences in cohort definition, outcome window, disease population, and preprocessing make direct cross-study comparison of point estimates unreliable. More broadly, the finding that additional modalities did not measurably improve on a strong structured-data baseline is consistent with a recurring theme in the multimodal clinical-prediction literature: the value an additional modality contributes depends on how much information it carries that is specifically relevant to the outcome of interest, not merely on its availability or on the sophistication of the fusion mechanism used to combine it.

### Calibration

The raw EHR-alone probe was systematically overconfident (calibration slope 0.12; Brier score 0.3203, worse than the 0.25 of a constant prediction of 0.5), whereas the fusion architectures produced more conservative probabilities (Table 8; Fig 2). This difference did not persist after recalibration: with a two-parameter logistic recalibration fitted on the validation set, EHR-alone reached a Brier score of 0.2388, comparable to the fusion models (0.2398–0.2439), with paired bootstrap differences that included zero. The raw gap therefore appears to reflect overfitting of the probability scale in a logistic probe trained on an 832-dimensional embedding with 895 patients, rather than a calibration benefit of multimodal fusion. Two further points limit clinical interpretation. First, all models remained only marginally better than a constant predictor (Brier skill 0.016–0.036), consistent with their modest discrimination (ROC-AUC 0.59–0.63). Second, because the cohort was enriched to 50% readmission and the models were trained with a class-weighted loss, none of the predicted probabilities, recalibrated or not, can be interpreted as absolute risks in the unselected COPD population (natural prevalence 18.1%) without recalibration to that population. Reporting calibration, and testing whether apparent calibration differences survive simple recalibration, should be part of multimodal prediction studies, since raw Brier scores can reward conservative probabilities rather than genuinely better models.

### Strengths

This study’s principal methodological strength is the systematic, empirically-driven audit of temporal data leakage across all four modality-linkage pipelines (see “Temporal alignment and leakage prevention” and Table 1), which identified three previously undetected issues, collectively affecting a substantial fraction of patients for two of the three affected modalities. Because each issue was identified through direct comparison against a verified ground-truth admission window rather than assumed absent, similar leakage may plausibly affect other studies using comparable MIMIC-IV-derived multimodal cohorts without an equally rigorous audit; we would encourage the adoption of comparable admission-window verification as standard practice in this area of research.

### Limitations

Several limitations should be considered when interpreting these findings. First, the outcome definition used here — 30-day, all-cause hospital readmission — captures readmission for any reason and is not restricted to readmissions attributable to a COPD exacerbation specifically (see “Outcome definition”). A more clinically specific outcome, defined using a validated administrative algorithm for identifying COPD exacerbations, may reveal a different balance of modality-specific signal than reported here, particularly for modalities such as chest radiography and clinical notes whose content may be more specifically informative for a respiratory-specific outcome than for hospital readmission in general; a follow-up study using such a validated, exacerbation-specific cohort, reusing the encoders, leakage-audited linkage procedures, and fusion architectures developed here, is planned.

Second, cohort size, and in particular per-modality coverage, constrained statistical power: the CXR modality’s coverage (13–16% across splits, reflecting genuine clinical practice rather than a pipeline artifact) limited both its unimodal evaluation (Table 4) and its non-linear adapter evaluation (Table 5, reported as inconclusive) to a small number of patients. The stratified analytic cohort used throughout this study (1,279 patients) represents a small fraction of the COPD patients available in the full MIMIC-IV database; re-deriving this study’s cohort at larger scale is a straightforward extension for future work.

Third, an asymmetry exists across the four modality encoders: the tabular EHR sub-encoder was itself trained in a supervised fashion against the study outcome before being frozen (see “Electronic health record (EHR)”), unlike the other three, purely self-supervised encoders. Although this does not constitute leakage — training, validation, and test splits were respected throughout — it may partly explain the EHR modality’s comparatively strong performance and should be considered when comparing modalities’ unimodal results.

Fourth, MIMIC-IV is a historical, single-health-system dataset; no external validation cohort was available for this study, and the generalizability of these findings to other institutions, populations, or time periods has not been established.

Fifth, the EHR-anchored residual fusion architecture was not independently hyperparameter-optimized (see “Training protocol and hyperparameter optimization”), unlike the naive concatenation and scalar gated fusion architectures, which received an identical and independently-optimized search; its results should be interpreted with this asymmetry in mind.

Sixth, the recalibration was fitted on the validation set that was also used for early stopping and hyperparameter selection of the fusion models, which may make their recalibration slightly optimistic; and, because the cohort was enriched to 50% readmission (natural prevalence 18.1%), the recalibrated probabilities do not apply to the unselected COPD population.

### Future directions

Beyond the exacerbation-specific follow-up study noted above, two further directions follow directly from these findings. First, given that a simple scalar gate did not outperform naive concatenation in this study, an architecture with more expressive cross-modal interactions, such as cross-modal attention, may be better positioned to recover an advantage from multimodal fusion in this already-compressed-embedding setting, and is planned as a subsequent study. Second, the temporal-proximity interpretation proposed above suggests that reformulating this task around a patient’s full admission history, or around the temporal proximity of each modality instance to the prediction point, rather than a single static index-admission snapshot, may prove a more productive direction than further refinement of the fusion mechanism alone.

## Conclusion

In a leakage-audited, MIMIC-IV-derived cohort of COPD patients, structured electronic health record (EHR) data alone predicted 30-day, all-cause hospital readmission as well as three independently-designed multimodal fusion architectures combining EHR data with ECG, chest radiograph, and clinical-note data. None of the tested fusion mechanisms provided a statistically significant improvement in discrimination over the EHR-alone baseline; the 95% confidence intervals for the difference in ROC-AUC (fusion minus EHR-alone) ranged from −0.13 to +0.07, so that gains larger than about 0.07 can be excluded but smaller ones cannot. A missingness-robustness stress test found that removing the auxiliary modalities improved, rather than degraded, fusion performance. The lower raw Brier scores of the fusion architectures did not persist after post-hoc recalibration, indicating that they reflected overconfidence of the EHR-alone probe rather than a benefit of fusion. These findings should be interpreted in light of the outcome definition used here (general readmission rather than a COPD-exacerbation-specific event), the cohort enrichment to 50% readmission (natural prevalence 18.1%), and the modest per-modality sample sizes available, all of which motivate the exacerbation-specific and larger-cohort follow-up studies described above. More broadly, we suggest that future studies combining multiple MIMIC-IV-derived data modalities budget explicit effort for empirical, admission-window-level auditing of modality-linkage pipelines, given the temporal-leakage issues identified here, which affected between 5% and 58% of patients depending on the modality.

## Acknowledgments

The authors thank the PhysioNet team and the MIMIC-IV data curators for making this dataset publicly available to credentialed researchers

## Supporting information

**S1 Table.** Full per-seed results for the three multimodal fusion architectures (naive concatenation, scalar gated fusion, EHR-anchored residual fusion), across validation and test splits.

| Model | Split | Seed | ROC-AUC | PR-AUC | Brier score |
| --- | --- | --- | --- | --- | --- |
| EHRAnchoredResidualFusion | Test | 0 | 0.5866 | 0.6452 | 0.2429 |
| EHRAnchoredResidualFusion | Test | 1 | 0.6356 | 0.6559 | 0.2385 |
| EHRAnchoredResidualFusion | Test | 2 | 0.5223 | 0.5467 | 0.2511 |
| EHRAnchoredResidualFusion | Test | 3 | 0.5866 | 0.6573 | 0.2423 |
| EHRAnchoredResidualFusion | Test | 4 | 0.5954 | 0.6816 | 0.2433 |
| NaiveConcatFusion | Test | 0 | 0.6355 | 0.6422 | 0.2549 |
| NaiveConcatFusion | Test | 1 | 0.6001 | 0.6205 | 0.2644 |
| NaiveConcatFusion | Test | 2 | 0.5619 | 0.5908 | 0.2574 |
| NaiveConcatFusion | Test | 3 | 0.6089 | 0.6338 | 0.2739 |
| NaiveConcatFusion | Test | 4 | 0.609 | 0.6294 | 0.2507 |
| ScalarGatedFusion | Test | 0 | 0.585 | 0.6309 | 0.2483 |
| ScalarGatedFusion | Test | 1 | 0.5979 | 0.6419 | 0.2413 |
| ScalarGatedFusion | Test | 2 | 0.5917 | 0.6311 | 0.2434 |
| ScalarGatedFusion | Test | 3 | 0.6128 | 0.6531 | 0.2456 |
| ScalarGatedFusion | Test | 4 | 0.5753 | 0.6121 | 0.2449 |
| EHRAnchoredResidualFusion | Validation | 0 | 0.6138 | 0.5785 | 0.2405 |
| EHRAnchoredResidualFusion | Validation | 1 | 0.6113 | 0.5759 | 0.242 |
| EHRAnchoredResidualFusion | Validation | 2 | 0.6474 | 0.6265 | 0.2397 |
| EHRAnchoredResidualFusion | Validation | 3 | 0.6079 | 0.5845 | 0.2421 |
| EHRAnchoredResidualFusion | Validation | 4 | 0.6192 | 0.5845 | 0.2408 |
| NaiveConcatFusion | Validation | 0 | 0.6272 | 0.5723 | 0.2573 |
| NaiveConcatFusion | Validation | 1 | 0.6308 | 0.5972 | 0.2496 |
| NaiveConcatFusion | Validation | 2 | 0.6306 | 0.5845 | 0.2448 |
| NaiveConcatFusion | Validation | 3 | 0.6114 | 0.5873 | 0.2575 |
| NaiveConcatFusion | Validation | 4 | 0.6163 | 0.5892 | 0.2533 |
| ScalarGatedFusion | Validation | 0 | 0.6221 | 0.5939 | 0.2517 |
| ScalarGatedFusion | Validation | 1 | 0.6135 | 0.5995 | 0.2457 |
| ScalarGatedFusion | Validation | 2 | 0.6079 | 0.5831 | 0.2456 |
| ScalarGatedFusion | Validation | 3 | 0.5937 | 0.57 | 0.2624 |
| ScalarGatedFusion | Validation | 4 | 0.6285 | 0.6075 | 0.2412 |

**S2 Table.** Full two-stage hyperparameter search results (Stage 1: shared projection dimensionality, learning rate, weight decay; Stage 2: dropout, batch size) for the naive concatenation and scalar gated fusion architectures, including all individually-searched configurations and their per-seed validation ROC-AUC.

| Model | Shared dim | Learning rate | Weight decay | Seed | Validation<br>ROC-AUC |
| --- | --- | --- | --- | --- | --- |
| NaiveConcatFusion | 32 | 0.0001 | 0.0001 | 0 | 0.6058 |
| NaiveConcatFusion | 32 | 0.0001 | 0.0001 | 1 | 0.6375 |
| NaiveConcatFusion | 32 | 0.0001 | 0.0001 | 2 | 0.6316 |
| NaiveConcatFusion | 32 | 0.0001 | 0.001 | 0 | 0.6201 |
| NaiveConcatFusion | 32 | 0.0001 | 0.001 | 1 | 0.6311 |
| NaiveConcatFusion | 32 | 0.0001 | 0.001 | 2 | 0.6312 |
| NaiveConcatFusion | 32 | 0.0001 | 0.01 | 0 | 0.6272 |
| NaiveConcatFusion | 32 | 0.0001 | 0.01 | 1 | 0.6308 |
| NaiveConcatFusion | 32 | 0.0001 | 0.01 | 2 | 0.6306 |
| NaiveConcatFusion | 32 | 0.0003 | 0.0001 | 0 | 0.6184 |
| NaiveConcatFusion | 32 | 0.0003 | 0.0001 | 1 | 0.6188 |
| NaiveConcatFusion | 32 | 0.0003 | 0.0001 | 2 | 0.6159 |
| NaiveConcatFusion | 32 | 0.0003 | 0.001 | 0 | 0.6431 |
| NaiveConcatFusion | 32 | 0.0003 | 0.001 | 1 | 0.6185 |
| NaiveConcatFusion | 32 | 0.0003 | 0.001 | 2 | 0.6186 |
| NaiveConcatFusion | 32 | 0.0003 | 0.01 | 0 | 0.605 |
| NaiveConcatFusion | 32 | 0.0003 | 0.01 | 1 | 0.6196 |
| NaiveConcatFusion | 32 | 0.0003 | 0.01 | 2 | 0.6245 |
| NaiveConcatFusion | 32 | 0.001 | 0.0001 | 0 | 0.6171 |
| NaiveConcatFusion | 32 | 0.001 | 0.0001 | 1 | 0.6281 |
| NaiveConcatFusion | 32 | 0.001 | 0.0001 | 2 | 0.6218 |
| NaiveConcatFusion | 32 | 0.001 | 0.001 | 0 | 0.6359 |
| NaiveConcatFusion | 32 | 0.001 | 0.001 | 1 | 0.6293 |
| NaiveConcatFusion | 32 | 0.001 | 0.001 | 2 | 0.6164 |
| NaiveConcatFusion | 32 | 0.001 | 0.01 | 0 | 0.6027 |
| NaiveConcatFusion | 32 | 0.001 | 0.01 | 1 | 0.634 |
| NaiveConcatFusion | 32 | 0.001 | 0.01 | 2 | 0.6144 |
| NaiveConcatFusion | 64 | 0.0001 | 0.0001 | 0 | 0.6089 |
| NaiveConcatFusion | 64 | 0.0001 | 0.0001 | 1 | 0.6155 |
| NaiveConcatFusion | 64 | 0.0001 | 0.0001 | 2 | 0.6205 |
| NaiveConcatFusion | 64 | 0.0001 | 0.001 | 0 | 0.6097 |
| NaiveConcatFusion | 64 | 0.0001 | 0.001 | 1 | 0.6132 |
| NaiveConcatFusion | 64 | 0.0001 | 0.001 | 2 | 0.6242 |
| NaiveConcatFusion | 64 | 0.0001 | 0.01 | 0 | 0.6093 |
| NaiveConcatFusion | 64 | 0.0001 | 0.01 | 1 | 0.6161 |
| NaiveConcatFusion | 64 | 0.0001 | 0.01 | 2 | 0.6399 |
| NaiveConcatFusion | 64 | 0.0003 | 0.0001 | 0 | 0.6082 |
| NaiveConcatFusion | 64 | 0.0003 | 0.0001 | 1 | 0.6205 |
| NaiveConcatFusion | 64 | 0.0003 | 0.0001 | 2 | 0.6241 |
| NaiveConcatFusion | 64 | 0.0003 | 0.001 | 0 | 0.606 |
| NaiveConcatFusion | 64 | 0.0003 | 0.001 | 1 | 0.6229 |
| NaiveConcatFusion | 64 | 0.0003 | 0.001 | 2 | 0.6245 |
| NaiveConcatFusion | 64 | 0.0003 | 0.01 | 0 | 0.6124 |
| NaiveConcatFusion | 64 | 0.0003 | 0.01 | 1 | 0.6031 |
| NaiveConcatFusion | 64 | 0.0003 | 0.01 | 2 | 0.6279 |
| NaiveConcatFusion | 64 | 0.001 | 0.0001 | 0 | 0.6146 |
| NaiveConcatFusion | 64 | 0.001 | 0.0001 | 1 | 0.6182 |
| NaiveConcatFusion | 64 | 0.001 | 0.0001 | 2 | 0.6167 |
| NaiveConcatFusion | 64 | 0.001 | 0.001 | 0 | 0.6158 |
| NaiveConcatFusion | 64 | 0.001 | 0.001 | 1 | 0.6349 |
| NaiveConcatFusion | 64 | 0.001 | 0.001 | 2 | 0.6264 |
| NaiveConcatFusion | 64 | 0.001 | 0.01 | 0 | 0.6115 |
| NaiveConcatFusion | 64 | 0.001 | 0.01 | 1 | 0.5913 |
| NaiveConcatFusion | 64 | 0.001 | 0.01 | 2 | 0.6007 |
| NaiveConcatFusion | 128 | 0.0001 | 0.0001 | 0 | 0.6179 |
| NaiveConcatFusion | 128 | 0.0001 | 0.0001 | 1 | 0.6192 |
| NaiveConcatFusion | 128 | 0.0001 | 0.0001 | 2 | 0.6327 |
| NaiveConcatFusion | 128 | 0.0001 | 0.001 | 0 | 0.6157 |
| NaiveConcatFusion | 128 | 0.0001 | 0.001 | 1 | 0.622 |
| NaiveConcatFusion | 128 | 0.0001 | 0.001 | 2 | 0.6311 |
| NaiveConcatFusion | 128 | 0.0001 | 0.01 | 0 | 0.6225 |
| NaiveConcatFusion | 128 | 0.0001 | 0.01 | 1 | 0.6243 |
| NaiveConcatFusion | 128 | 0.0001 | 0.01 | 2 | 0.6352 |
| NaiveConcatFusion | 128 | 0.0003 | 0.0001 | 0 | 0.6139 |
| NaiveConcatFusion | 128 | 0.0003 | 0.0001 | 1 | 0.6202 |
| NaiveConcatFusion | 128 | 0.0003 | 0.0001 | 2 | 0.6313 |
| NaiveConcatFusion | 128 | 0.0003 | 0.001 | 0 | 0.6148 |
| NaiveConcatFusion | 128 | 0.0003 | 0.001 | 1 | 0.6189 |
| NaiveConcatFusion | 128 | 0.0003 | 0.001 | 2 | 0.6318 |
| NaiveConcatFusion | 128 | 0.0003 | 0.01 | 0 | 0.614 |
| NaiveConcatFusion | 128 | 0.0003 | 0.01 | 1 | 0.6252 |
| NaiveConcatFusion | 128 | 0.0003 | 0.01 | 2 | 0.6332 |
| NaiveConcatFusion | 128 | 0.001 | 0.0001 | 0 | 0.622 |
| NaiveConcatFusion | 128 | 0.001 | 0.0001 | 1 | 0.6234 |
| NaiveConcatFusion | 128 | 0.001 | 0.0001 | 2 | 0.6275 |
| NaiveConcatFusion | 128 | 0.001 | 0.001 | 0 | 0.6228 |
| NaiveConcatFusion | 128 | 0.001 | 0.001 | 1 | 0.6219 |
| NaiveConcatFusion | 128 | 0.001 | 0.001 | 2 | 0.6295 |
| NaiveConcatFusion | 128 | 0.001 | 0.01 | 0 | 0.6182 |
| NaiveConcatFusion | 128 | 0.001 | 0.01 | 1 | 0.6011 |
| NaiveConcatFusion | 128 | 0.001 | 0.01 | 2 | 0.6355 |
| ScalarGatedFusion | 32 | 0.0001 | 0.0001 | 0 | 0.5936 |
| ScalarGatedFusion | 32 | 0.0001 | 0.0001 | 1 | 0.5961 |
| ScalarGatedFusion | 32 | 0.0001 | 0.0001 | 2 | 0.5849 |
| ScalarGatedFusion | 32 | 0.0001 | 0.001 | 0 | 0.5957 |
| ScalarGatedFusion | 32 | 0.0001 | 0.001 | 1 | 0.6034 |
| ScalarGatedFusion | 32 | 0.0001 | 0.001 | 2 | 0.5843 |
| ScalarGatedFusion | 32 | 0.0001 | 0.01 | 0 | 0.5904 |
| ScalarGatedFusion | 32 | 0.0001 | 0.01 | 1 | 0.5973 |
| ScalarGatedFusion | 32 | 0.0001 | 0.01 | 2 | 0.5882 |
| ScalarGatedFusion | 32 | 0.0003 | 0.0001 | 0 | 0.6105 |
| ScalarGatedFusion | 32 | 0.0003 | 0.0001 | 1 | 0.6025 |
| ScalarGatedFusion | 32 | 0.0003 | 0.0001 | 2 | 0.6018 |
| ScalarGatedFusion | 32 | 0.0003 | 0.001 | 0 | 0.6076 |
| ScalarGatedFusion | 32 | 0.0003 | 0.001 | 1 | 0.6033 |
| ScalarGatedFusion | 32 | 0.0003 | 0.001 | 2 | 0.6032 |
| ScalarGatedFusion | 32 | 0.0003 | 0.01 | 0 | 0.608 |
| ScalarGatedFusion | 32 | 0.0003 | 0.01 | 1 | 0.6023 |
| ScalarGatedFusion | 32 | 0.0003 | 0.01 | 2 | 0.6002 |
| ScalarGatedFusion | 32 | 0.001 | 0.0001 | 0 | 0.6067 |
| ScalarGatedFusion | 32 | 0.001 | 0.0001 | 1 | 0.612 |
| ScalarGatedFusion | 32 | 0.001 | 0.0001 | 2 | 0.6218 |
| ScalarGatedFusion | 32 | 0.001 | 0.001 | 0 | 0.6077 |
| ScalarGatedFusion | 32 | 0.001 | 0.001 | 1 | 0.6107 |
| ScalarGatedFusion | 32 | 0.001 | 0.001 | 2 | 0.6219 |
| ScalarGatedFusion | 32 | 0.001 | 0.01 | 0 | 0.6033 |
| ScalarGatedFusion | 32 | 0.001 | 0.01 | 1 | 0.6036 |
| ScalarGatedFusion | 32 | 0.001 | 0.01 | 2 | 0.6181 |
| ScalarGatedFusion | 64 | 0.0001 | 0.0001 | 0 | 0.5933 |
| ScalarGatedFusion | 64 | 0.0001 | 0.0001 | 1 | 0.587 |
| ScalarGatedFusion | 64 | 0.0001 | 0.0001 | 2 | 0.6158 |
| ScalarGatedFusion | 64 | 0.0001 | 0.001 | 0 | 0.5925 |
| ScalarGatedFusion | 64 | 0.0001 | 0.001 | 1 | 0.5886 |
| ScalarGatedFusion | 64 | 0.0001 | 0.001 | 2 | 0.6148 |
| ScalarGatedFusion | 64 | 0.0001 | 0.01 | 0 | 0.5921 |
| ScalarGatedFusion | 64 | 0.0001 | 0.01 | 1 | 0.5884 |
| ScalarGatedFusion | 64 | 0.0001 | 0.01 | 2 | 0.6202 |
| ScalarGatedFusion | 64 | 0.0003 | 0.0001 | 0 | 0.5852 |
| ScalarGatedFusion | 64 | 0.0003 | 0.0001 | 1 | 0.5981 |
| ScalarGatedFusion | 64 | 0.0003 | 0.0001 | 2 | 0.6183 |
| ScalarGatedFusion | 64 | 0.0003 | 0.001 | 0 | 0.5982 |
| ScalarGatedFusion | 64 | 0.0003 | 0.001 | 1 | 0.5971 |
| ScalarGatedFusion | 64 | 0.0003 | 0.001 | 2 | 0.6191 |
| ScalarGatedFusion | 64 | 0.0003 | 0.01 | 0 | 0.596 |
| ScalarGatedFusion | 64 | 0.0003 | 0.01 | 1 | 0.5999 |
| ScalarGatedFusion | 64 | 0.0003 | 0.01 | 2 | 0.6159 |
| ScalarGatedFusion | 64 | 0.001 | 0.0001 | 0 | 0.5994 |
| ScalarGatedFusion | 64 | 0.001 | 0.0001 | 1 | 0.6058 |
| ScalarGatedFusion | 64 | 0.001 | 0.0001 | 2 | 0.6198 |
| ScalarGatedFusion | 64 | 0.001 | 0.001 | 0 | 0.5912 |
| ScalarGatedFusion | 64 | 0.001 | 0.001 | 1 | 0.6073 |
| ScalarGatedFusion | 64 | 0.001 | 0.001 | 2 | 0.6188 |
| ScalarGatedFusion | 64 | 0.001 | 0.01 | 0 | 0.603 |
| ScalarGatedFusion | 64 | 0.001 | 0.01 | 1 | 0.6101 |
| ScalarGatedFusion | 64 | 0.001 | 0.01 | 2 | 0.6188 |
| ScalarGatedFusion | 128 | 0.0001 | 0.0001 | 0 | 0.5858 |
| ScalarGatedFusion | 128 | 0.0001 | 0.0001 | 1 | 0.599 |
| ScalarGatedFusion | 128 | 0.0001 | 0.0001 | 2 | 0.6022 |
| ScalarGatedFusion | 128 | 0.0001 | 0.001 | 0 | 0.586 |
| ScalarGatedFusion | 128 | 0.0001 | 0.001 | 1 | 0.5979 |
| ScalarGatedFusion | 128 | 0.0001 | 0.001 | 2 | 0.6033 |
| ScalarGatedFusion | 128 | 0.0001 | 0.01 | 0 | 0.5813 |
| ScalarGatedFusion | 128 | 0.0001 | 0.01 | 1 | 0.5978 |
| ScalarGatedFusion | 128 | 0.0001 | 0.01 | 2 | 0.6101 |
| ScalarGatedFusion | 128 | 0.0003 | 0.0001 | 0 | 0.5954 |
| ScalarGatedFusion | 128 | 0.0003 | 0.0001 | 1 | 0.6035 |
| ScalarGatedFusion | 128 | 0.0003 | 0.0001 | 2 | 0.6159 |
| ScalarGatedFusion | 128 | 0.0003 | 0.001 | 0 | 0.5949 |
| ScalarGatedFusion | 128 | 0.0003 | 0.001 | 1 | 0.6027 |
| ScalarGatedFusion | 128 | 0.0003 | 0.001 | 2 | 0.617 |
| ScalarGatedFusion | 128 | 0.0003 | 0.01 | 0 | 0.5982 |
| ScalarGatedFusion | 128 | 0.0003 | 0.01 | 1 | 0.6012 |
| ScalarGatedFusion | 128 | 0.0003 | 0.01 | 2 | 0.617 |
| ScalarGatedFusion | 128 | 0.001 | 0.0001 | 0 | 0.6041 |
| ScalarGatedFusion | 128 | 0.001 | 0.0001 | 1 | 0.6016 |
| ScalarGatedFusion | 128 | 0.001 | 0.0001 | 2 | 0.6171 |
| ScalarGatedFusion | 128 | 0.001 | 0.001 | 0 | 0.6052 |
| ScalarGatedFusion | 128 | 0.001 | 0.001 | 1 | 0.6018 |
| ScalarGatedFusion | 128 | 0.001 | 0.001 | 2 | 0.6195 |
| ScalarGatedFusion | 128 | 0.001 | 0.01 | 0 | 0.6054 |
| ScalarGatedFusion | 128 | 0.001 | 0.01 | 1 | 0.5932 |
| ScalarGatedFusion | 128 | 0.001 | 0.01 | 2 | 0.6206 |

Stage 2
| Model | Dropout | Batch size | Seed | Validation<br>ROC-AUC | Fixed<br>shared dim<br>(from Stage<br>1) | Fixed<br>learning<br>rate (from<br>Stage 1) | Fixed weight<br>decay (from<br>Stage 1) |
| --- | --- | --- | --- | --- | --- | --- | --- |
| NaiveConcatFusion | 0.1 | 8 | 0 | 0.6203 | 32 | 0.0001 | 0.01 |
| NaiveConcatFusion | 0.1 | 8 | 1 | 0.6306 | 32 | 0.0001 | 0.01 |
| NaiveConcatFusion | 0.1 | 8 | 2 | 0.6235 | 32 | 0.0001 | 0.01 |
| NaiveConcatFusion | 0.1 | 16 | 0 | 0.6244 | 32 | 0.0001 | 0.01 |
| NaiveConcatFusion | 0.1 | 16 | 1 | 0.6291 | 32 | 0.0001 | 0.01 |
| NaiveConcatFusion | 0.1 | 16 | 2 | 0.6345 | 32 | 0.0001 | 0.01 |
| NaiveConcatFusion | 0.1 | 32 | 0 | 0.6042 | 32 | 0.0001 | 0.01 |
| NaiveConcatFusion | 0.1 | 32 | 1 | 0.6351 | 32 | 0.0001 | 0.01 |
| NaiveConcatFusion | 0.1 | 32 | 2 | 0.6201 | 32 | 0.0001 | 0.01 |
| NaiveConcatFusion | 0.2 | 8 | 0 | 0.6135 | 32 | 0.0001 | 0.01 |
| NaiveConcatFusion | 0.2 | 8 | 1 | 0.627 | 32 | 0.0001 | 0.01 |
| NaiveConcatFusion | 0.2 | 8 | 2 | 0.6263 | 32 | 0.0001 | 0.01 |
| NaiveConcatFusion | 0.2 | 16 | 0 | 0.6272 | 32 | 0.0001 | 0.01 |
| NaiveConcatFusion | 0.2 | 16 | 1 | 0.6308 | 32 | 0.0001 | 0.01 |
| NaiveConcatFusion | 0.2 | 16 | 2 | 0.6306 | 32 | 0.0001 | 0.01 |
| NaiveConcatFusion | 0.2 | 32 | 0 | 0.6042 | 32 | 0.0001 | 0.01 |
| NaiveConcatFusion | 0.2 | 32 | 1 | 0.6336 | 32 | 0.0001 | 0.01 |
| NaiveConcatFusion | 0.2 | 32 | 2 | 0.6339 | 32 | 0.0001 | 0.01 |
| NaiveConcatFusion | 0.3 | 8 | 0 | 0.6102 | 32 | 0.0001 | 0.01 |
| NaiveConcatFusion | 0.3 | 8 | 1 | 0.6201 | 32 | 0.0001 | 0.01 |
| NaiveConcatFusion | 0.3 | 8 | 2 | 0.6281 | 32 | 0.0001 | 0.01 |
| NaiveConcatFusion | 0.3 | 16 | 0 | 0.6089 | 32 | 0.0001 | 0.01 |
| NaiveConcatFusion | 0.3 | 16 | 1 | 0.6264 | 32 | 0.0001 | 0.01 |
| NaiveConcatFusion | 0.3 | 16 | 2 | 0.6341 | 32 | 0.0001 | 0.01 |
| NaiveConcatFusion | 0.3 | 32 | 0 | 0.6126 | 32 | 0.0001 | 0.01 |
| NaiveConcatFusion | 0.3 | 32 | 1 | 0.6267 | 32 | 0.0001 | 0.01 |
| NaiveConcatFusion | 0.3 | 32 | 2 | 0.6093 | 32 | 0.0001 | 0.01 |
| ScalarGatedFusion | 0.1 | 8 | 0 | 0.596 | 32 | 0.001 | 0.0001 |
| ScalarGatedFusion | 0.1 | 8 | 1 | 0.6206 | 32 | 0.001 | 0.0001 |
| ScalarGatedFusion | 0.1 | 8 | 2 | 0.6054 | 32 | 0.001 | 0.0001 |
| ScalarGatedFusion | 0.1 | 16 | 0 | 0.6175 | 32 | 0.001 | 0.0001 |
| ScalarGatedFusion | 0.1 | 16 | 1 | 0.5998 | 32 | 0.001 | 0.0001 |
| ScalarGatedFusion | 0.1 | 16 | 2 | 0.6151 | 32 | 0.001 | 0.0001 |
| ScalarGatedFusion | 0.1 | 32 | 0 | 0.6239 | 32 | 0.001 | 0.0001 |
| ScalarGatedFusion | 0.1 | 32 | 1 | 0.6238 | 32 | 0.001 | 0.0001 |
| ScalarGatedFusion | 0.1 | 32 | 2 | 0.607 | 32 | 0.001 | 0.0001 |
| ScalarGatedFusion | 0.2 | 8 | 0 | 0.5985 | 32 | 0.001 | 0.0001 |
| ScalarGatedFusion | 0.2 | 8 | 1 | 0.6049 | 32 | 0.001 | 0.0001 |
| ScalarGatedFusion | 0.2 | 8 | 2 | 0.5988 | 32 | 0.001 | 0.0001 |
| ScalarGatedFusion | 0.2 | 16 | 0 | 0.6067 | 32 | 0.001 | 0.0001 |
| ScalarGatedFusion | 0.2 | 16 | 1 | 0.612 | 32 | 0.001 | 0.0001 |
| ScalarGatedFusion | 0.2 | 16 | 2 | 0.6218 | 32 | 0.001 | 0.0001 |
| ScalarGatedFusion | 0.2 | 32 | 0 | 0.6268 | 32 | 0.001 | 0.0001 |
| ScalarGatedFusion | 0.2 | 32 | 1 | 0.6141 | 32 | 0.001 | 0.0001 |
| ScalarGatedFusion | 0.2 | 32 | 2 | 0.6058 | 32 | 0.001 | 0.0001 |
| ScalarGatedFusion | 0.3 | 8 | 0 | 0.5978 | 32 | 0.001 | 0.0001 |
| ScalarGatedFusion | 0.3 | 8 | 1 | 0.6061 | 32 | 0.001 | 0.0001 |
| ScalarGatedFusion | 0.3 | 8 | 2 | 0.5968 | 32 | 0.001 | 0.0001 |
| ScalarGatedFusion | 0.3 | 16 | 0 | 0.6011 | 32 | 0.001 | 0.0001 |
| ScalarGatedFusion | 0.3 | 16 | 1 | 0.6049 | 32 | 0.001 | 0.0001 |
| ScalarGatedFusion | 0.3 | 16 | 2 | 0.6149 | 32 | 0.001 | 0.0001 |
| ScalarGatedFusion | 0.3 | 32 | 0 | 0.6332 | 32 | 0.001 | 0.0001 |
| ScalarGatedFusion | 0.3 | 32 | 1 | 0.6005 | 32 | 0.001 | 0.0001 |
| ScalarGatedFusion | 0.3 | 32 | 2 | 0.6041 | 32 | 0.001 | 0.0001 |

## Notes

### Competing Interest Statement

The authors have declared no competing interest.

### Author Declarations

The Institutional Review Board of Beth Israel Deaconess Medical Center (Boston, Massachusetts, USA) approved the creation and sharing of the MIMIC-IV database and waived the requirement for informed consent. This work is a secondary analysis of this de-identified database, accessed under the PhysioNet Credentialed Health Data Use Agreement.

